# Optimal Control and Cost-effectiveness Analysis of an HPV-Chlamydia Trachomatis co-infection model

**DOI:** 10.1101/2020.09.07.20190025

**Authors:** A. Omame, C. U. Nnanna, S. C. Inyama

## Abstract

In this work, a co-infection model for human papillomavirus (HPV) and Chlamydia trachomatis with cost-effectiveness optimal control analysis is developed and analyzed. The disease-free equilibrium of the co-infection model is shown not to be globally asymptotically stable, when the associated reproduction number is less unity. It is proven that the model undergoes the phenomenon of backward bifurcation when the associated reproduction number is less than unity. It is also shown that HPV re-infection (*ε*_p_ ≠ 0) induced the phenomenon of backward bifurcation. Numerical simulations of the optimal control model showed that: (i) focusing on HPV intervention strategy alone (HPV prevention and screening), in the absence of Chlamydia trachomatis control, leads to a positive population level impact on the total number of individuals singly infected with Chlamydia trachomatis, (ii) Concentrating on Chlamydia trachomatis intervention controls alone (Chlamydia trachomatis prevention and treatment), in the absence of HPV intervention strategies, a positive population level impact is observed on the total number of individuals singly infected with HPV. Moreover, the strategy that combines and implements HPV and Chlamydia trachomatis prevention controls is the most cost-effective of all the control strategies in combating the co-infections of HPV and Chlamydia trachomatis.

## 1 Introduction

Chlamydia trachomatis is one of the most common curable bacterial sexually transmitted infections (STIs) [6]. According to the World Health Organization (WHO), an estimated 50 million women are infected with Chlamydia trachomatis around the world, with Southern Asia and Sub-Saharan Africa recording 34 million cases [12]. In the United states, Chlamydia trachomatis is the most common reported bacterial STI. According to the Centers for Disease Control and Prevention (CDC), about 1, 758, 668 new cases of Chlamydia trachomatis were reported in the United States in 2018. Chlamydia trachomatis infection is curable with antibiotics [6]. Untreated Chlamydia trachomatis infection can result in up to 40% cases of pelvic inflammatory disease (PID) and subsequent infertility, especially in women [12]. Other consequences of untreated or complicated Chlamydia trachomatis infection include ectopic pregnancy and chronic pelvic pain [20]. Human papillomavirus (HPV) infection is one of the most common STIs [47]. Although most infections do not cause illness, persistent HPV infection is a necessary cause of cervical cancer, anal cancer and other forms of cancer [47]. In 2018, an estimated 311, 000 women died of cervical cancer, with more than 80% of these recorded deaths occuring in less developed countries in Asia and Sub-Saharan Africa [47].

Substantial cervical cancer intervention strategies include prevention of incident infection through HPV vaccination and condom use, screening and treatment of pre-cancerous lesions, diagnosis and treatment of invasive cervical cancer, as well as palliative care for fully developed cervical cancer patients [47]. Vaccines which can prevent incident infection with cancer-causing HPV types have been recommended by the World Health Organization (WHO) and are currently being administered in many countries [47]. They include: the bivalent *Cervarix* vaaccine, the quadrivalent *Gardasil 4* vaccine and the newly introduced *Gardasil 9* vaccine. Cervical cancer screening testing involves testing for pre-cancer and cancer among individuals who display no symptoms and may not even feel sick [47]. When screening detects pre-cancerous lesions, these can be easily treated and cancer can be avoided. Presently, the WHO has recommended three types of screening tests: HPV testing for high risk HPV types, visual inspection with *Acetic Acid* (VIA) as well as conventional (Pap) test and liquid-based cytology (LBC) [47]. Also, the WHO recommends the use of cryotherapy and Loop Electrosurgical Excision Procedure (LEEP) for treatment of pre-cancer lesions.

Co-infection between HPV and Chlamydia trachomatis have been well explored epidemiologically [10, 16, 19, 33, 38, 46]. Nonato *et al*. [19] studied the interaction between HPV and Chlamydia trachomatis and showed that the co-infection of the two diseases can increase progression to cervical neoplasia by women. In another paper, Samoff *et al*. [33] opined that Chlamydia trachomatis was associated with persistent HPV infection. Clinical studies have equally shown that women infected with Chlamydia trachomatis have a higher risk of developing cervical cancer than uninfected women [46]. In another epidemiological study, it was revealed that women with Chlamydia trachomatis have an increased susceptibility to persistent HPV infection in comparison to Chlamydia trachomatis uninfected women [35]. Moreover, Seraceni *et al*.[34] pointed out that younger women infected with chronic Chlamydia trachomatis infection have a higher risk of infection with multiple HPV types. In addition, Ssedyabane *et al*. [38] conducted a pilot study to investigate the co-infection between HPV and Chlamydia trachomatis in Mbarara Region in Uganda. The authors showed that there is a strong correlation between the co-infection of the two diseases and cervical intraepithelial neoplasia (CIN). Chlamydia trachomatis infection can lead to chronic inflammation, cervical hypotrophy and squamous metaplasia, which are potential target cells for HPV infection [29]. Furthermore, a higher prevalence of Chalmydia trachomatis DNA or *IgG* antibodies have been observed in HPV positve samples in comparison to HPV negative samples, indicating a strong correlation between the two diseases [10, 16]. Both Chlamydia trachomatis and HPV infections may increase the expression of *Ki67*, which is strongly linked with the proliferation and growth of squamous cell [39].

Mathematical modelling has been used extensively in studying the behaviour of infectious diseases, including their co-infections [1, 7, 13, 18, 23, 28, 41, 43, 44]. Particularly, Several mathematical models have been developed to understand the transmission dyanamics of Chlamydia trachomatis infections. Sharomi and Gumel [36] developed a two-sex comprehensive mathematical model to assess the impact of treatment on the dynamics of Chlamydia trachomatis. Their model exhibited the phenomenon of backward bifurcation caused by the re-infection of individuals who had recovered from a previous infection with the disease. Simulations of the model in [36] showed that the implementation of Chlamydia treatment strategy for only males or only females could significantly save new cases of Chlamydia infection in the opposite sex. In another paper, Sharomi and Gumel [37] developed a risk-structured, two-group deterministic model for Chlamydia trachomatis, which stratified the population based on risk of acquiring or transmitting infection. They showed that stratifying the Chlamydia trachomatis model, based on the risk of acquiring or transmitting infection, induced the phenomenon of backward bifurcation even when the re-infection of recovered individuals did not occur. Samanta [32] developed a mathematical model for Chlamydia with pulse vacciantion strategy. He showed using simulations with MATLAB, the conditions under which the disease will go into extinction and when the disease will persist in the population. For a detailed review of HPV models in the literature, see the work of Omame *et al*. [25] and the references therein. Recently, optimal control and cost-effectiveness analysis have been applied to deterministic mathematical models [17, 21, 31, 40]. Although numerous epidemiological evidences to support the co-infection of HPV and Chlamydia trachomatis exist in the literature, no robust optimal control mathematical model has been developed to better understand the dynamics of the co-infection of the two diseases. Hence, it is appropriate to study the optimal control and cost-effectiveness analysis of the co-infections of HPV and Chlamydia trachomatis.

This paper assesses the impact of HPV screening, Chlamydia trachomatis treatment as well as preventive strategies for both diseases on the control and management of their co-infections, and subsequent prevention of cancers and pelvic inflammatory disease (PID), using a mathematical model. Optimal control and cost-effectiveness analysis is also carried out on the model to determine the most cost-effective strategy in combating the co-infection of both diseases. To the best of the authors’ knowledege, a co-infection model for HPV and Chlamydia trachomatis is being considered for the first time.

The rest of the paper is organized as follows: The model formulation and basic properties are discussed in Section 2. The co-infection model without controls is analyzed qualitatively in Section 3. The optimal control model is considered in Section 4, simulations of the model are carried out in Section 5, while Section 6 gives the concluding remarks.

## 2 Model formulation

The total sexually active population at time *t*, denoted by *N*_H_(*t*), is divided into eight mutually-exclusive compartments: Susceptible individuals (*S*_H_(*t*)), individuals infected with HPV (*I*_HP_(*t*)), infectious individuals screened for HPV infection (*I*_SHP_(*t*)), individuals who have recovered from or cleared of HPV infection (*R*_HP_(*t*)), individuals infected with Chlamydia trachomatis (*I*_CL_(*t*)), individuals who have recovered from Chlamydia trachomatis infection (*R*_CL_(*t*)), individuals dually infected with HPV and Chlamydia trachomatis (*I*_HPCL_(*t*)), infectious individuals screened for HPV infection and infected with Chlamydia trachomatis (*I*_SHPCL_). Therefore,

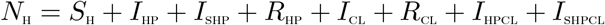

The population of sexually active individuals, *S*_H_, is generated by the recruitment of individuals at a rate Λ_H_. This population is decreased upon infection with HPV, following effective contact with both singly and dually infected individuals with HPV at the rate:

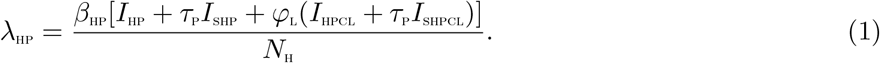

The population of sexually active individuals, *S*_H_, is also decreased upon infection with Chlamydia trachomatis, acquired due to effective contact with both singly and dually infected individuals with Chlamydia trachomatis at the rate:

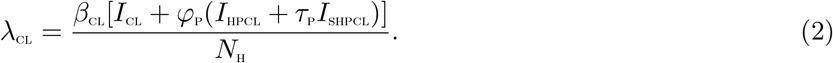

The modification parameter 0 *< τ*_P_ < 1, accounts for reduced probability of transmission by infectious individuals screened for HPV infection (that is, the parameter *τ*_P_ measures the efficacy of screening in reducing the risk of transmission of HPV). It is assumed that individuals screened for HPV infection report early for treatment, thereby reducing their transmission probability. Their risk of dying as a result of the disease is also assumed negligible. The parameter *φ*_L_(*φ*_L_ *≥* 1) is a modification term accounting for the increased infectiousness of dually infected individuals due to Chlamaydia trachomatis infection. Similarly, the parameter *φ*_P_(*φ*_P_ *≥* 1) accounts for increased infectiousness of dually infected individuals due to HPV infection. This population is further reduced by natural death (at a rate *μ*_H_. Natural death occurs in all the epidemiological compartments at this rate). In (1), *β*_HP_ is the effective contact rate for the transmission of HPV infection. Likewise, in (2), *β*_CL_ denotes the effective contact rate for the transmission of Chlamydia trachomatis infection. It is asumed in the model that individuals infected with Chlamydia trachomatis have an increased susceptibility to infection with HPV [19, 34, 35]. Likewise, individuals with HPV infection have higher chances of getting infected with Chlamydia trachomatis [10, 16].

Based on the above formulations and assumptions, the HPV-Chlamydia trachomatis co-infection model is given by the following system of deterministic differential equations (the flow diagram of the model is shown in Figure 1 and the associated parameters of the model are presented in Table 2):

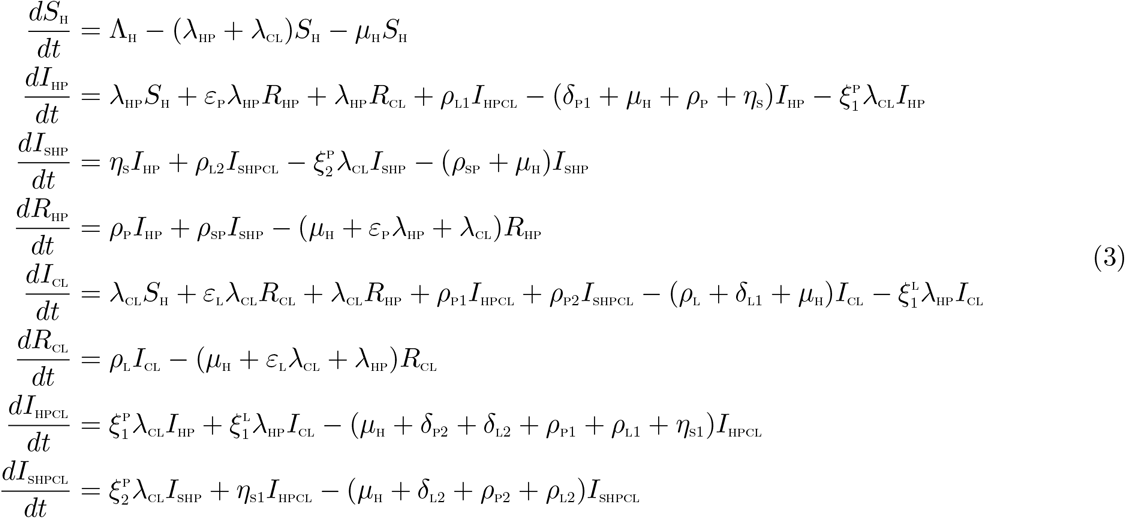

**Figure 1:**
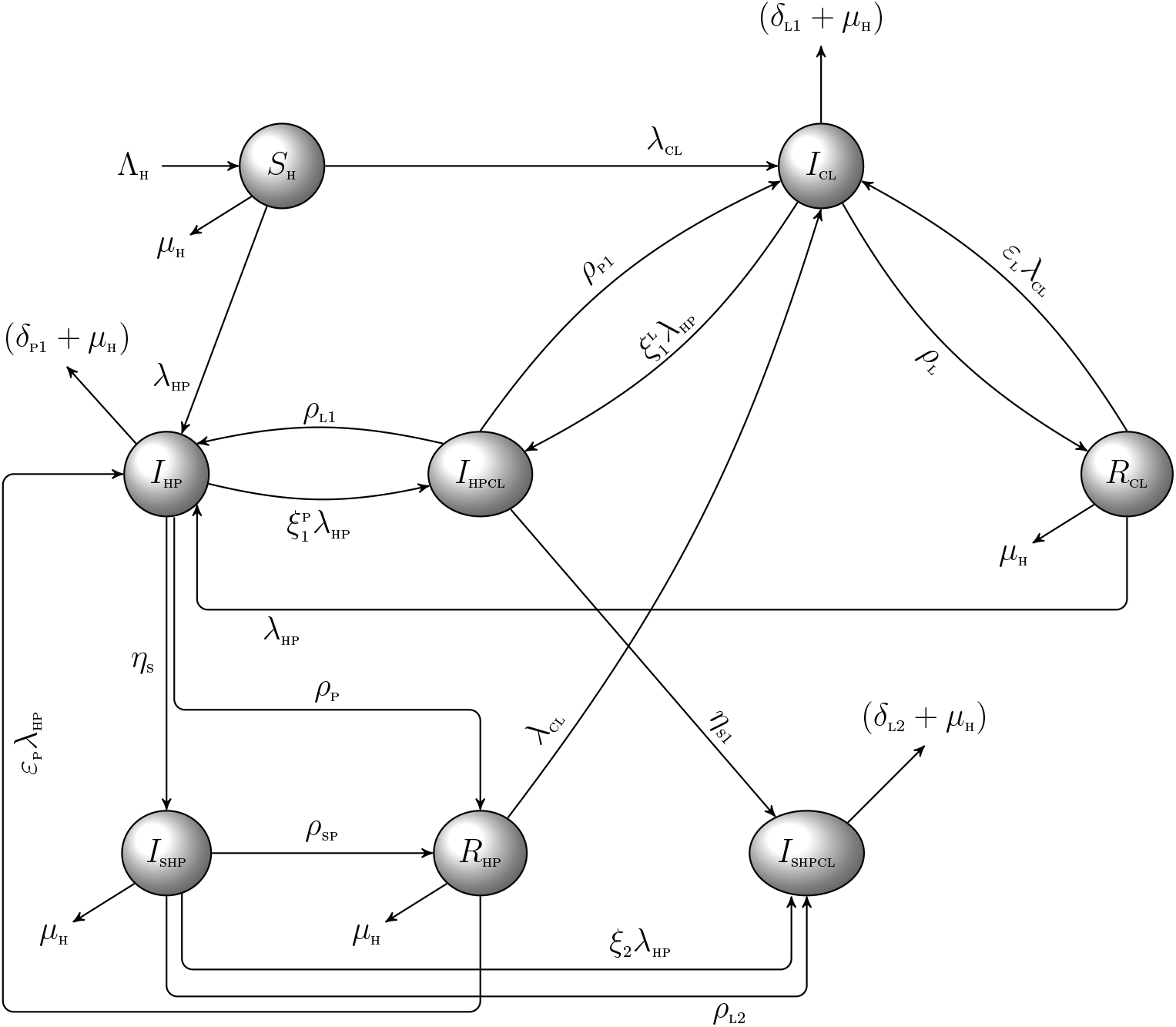
Schematic diagram of the model (3)

**Table 1:** PRCC values for the HPV-Chlamydia trachomatis co-infection model (3) parameters using the total number of singly infected individuals: HPV (*I*_HP_), Chlamydia trachomatis (*I*_CL_), and individuals dually infected with HPV and Chlamydia trachomatis (*I*_HPCL_), respectively, as well as the associated reproduction numbers for HPV, 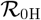, and Chlamydia trachomatis, 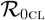, respectively, as response functions. Paramters which strongly influence the dynamics of the co-infection model with respect to each of the response functions are shown in bold fonts.

| Parameters | $I_{\text{HP}}$ | $I_{\text{CL}}$ | $I_{\text{HPCL}}$ | $\mathcal{R}_{0\text{HP}}$ | $\mathcal{R}_{0\text{CL}}$ |
| --- | --- | --- | --- | --- | --- |
| $\mu_{\text{H}}$ | -0.0907 | -0.1446 | -0.2267 | -0.2966 | -0.2768 |
| $\beta_{\text{HP}}$ | **0.7144** | **0.4504** | **0.7352** | **0.9109** | — |
| $\beta_{\text{CL}}$ | -0.3704 | **0.7686** | **0.7772** | — | **0.9333** |
| $\varepsilon_{\text{P}}$ | 0.1187 | 0.0081 | 0.00378 | — | — |
| $\varepsilon_{\text{L}}$ | -0.0411 | 0.1390 | 0.0235 | — | — |
| $\eta_{\text{S}}$ | -0.1697 | 0.0241 | -0.1123 | -0.2972 | — |
| $\eta_{\text{S}1}$ | -0.0992 | -0.0660 | **-0.4016** | — | — |
| $\rho_{\text{P}}$ | -0.2339 | 0.0351 | -0.1122 | **-0.8833** | — |
| $\rho_{\text{SP}}$ | 0.0013 | 0.0716 | 0.0180 | **-0.5813** | — |
| $\rho_{\text{P}1}$ | -0.0780 | **0.4483** | -0.2613 | — | — |
| $\rho_{\text{P}2}$ | 0.0661 | 0.1218 | 0.2024 | — | — |
| $\rho_{\text{L}}$ | 0.0254 | -0.3618 | -0.1380 | — | **-0.9548** |
| $\rho_{\text{L}1}$ | **0.5196** | -0.1482 | -0.3858 | — | — |
| $\rho_{\text{L}2}$ | 0.0062 | -0.0287 | -0.0278 | — | — |
| $\delta_{\text{L}1}$ | -0.0056 | -0.0376 | 0.0054 | — | -0.1198 |
| $\delta_{\text{L}2}$ | -0.0493 | -0.0539 | -0.398 | — | — |
| $\delta_{\text{P}1}$ | 0.0173 | 0.0090 | -0.0230 | -0.0481 | — |
| $\delta_{\text{P}2}$ | 0.0198 | -0.0165 | 0.058 | — | — |
| $\varphi_{\text{P}}$ | -0.2266 | 0.3749 | **0.5488** | — | — |
| $\varphi_{\text{L}}$ | 0.3460 | -0.3086 | **0.5124** | — | — |
| $\tau_{\text{P}}$ | 0.1251 | 0.0711 | **0.4002** | **0.5518** | — |
| $\xi_1^{\text{P}}$ | -0.2933 | 0.1845 | **0.4017** | — | — |
| $\xi_2^{\text{P}}$ | 0.000665 | 0.1103 | 0.0608 | — | — |
| $\xi_1^{\text{L}}$ | 0.1739 | -0.3048 | **0.4203** | — | — |

**Table 2:** Description of parameters in the model (3)

| Parameter | Description | Value | Reference |
| --- | --- | --- | --- |
| $\Lambda_H$ | Recruitment rate | 365,052 | [42] |
| $\mu_H$ | Natural death rate | 0.0178 | [42] |
| $\beta_{HP}$ | Effective contact rate for HPV transmission | 1.0 | [27] |
| $\beta_{CL}$ | Effective contact rate for Chlamydia trachomatis transmission | 1.1 | [36] |
| $\tau_P$ | modification parameter accounting for reduced transmission | | |
|  | probability of HPV infected individuals who have undergone screening, relative to those who have not been screened | 0.9 | Assumed |
| $\delta_{P1}, \delta_{P2}$ | HPV-induced death rate for singly and dually infected individuals, respectively | 0.001 | [26] |
| $\delta_{L1}, \delta_{L2}$ | Chlamydia trachomatis-induced death rate for singly and dually infected individuals, respectively | 0.05 | Assumed |
| $\varepsilon_P$ | HPV re-infection rate for individuals who recovered from HPV | 0.3 | [25] |
| $\varepsilon_L$ | Chlamydia trachomatis re-infection rate for individuals who recovered from previous infection | 0.3 | [36] |
| $\varphi_L$ | modification parameter accounting for increased transmission | | |
|  | probability of dually infected individuals due to Chlamydia trachomatis | 1.3 | Assumed |
| $\varphi_P$ | modification parameter accounting for increased transmission | | |
|  | probability of dually infected individuals due to HPV | 1.3 | [26] |
| $\xi_1^P, \xi_2^P$ | Modification parameter for increased susceptibility to Chlamydia trachomatis due to HPV infection | 1.3 | [34, 35] |
| $\xi_1^L$ | Modification parameter accounting for increased susceptibility to HPV due to Chlamydia trachomatis infection | 1.2 | [10, 16] |
| $\eta_S, \eta_{S1}$ | Screening rate for HPV infected individuals | 0.90 | Assumed |
| $\rho_P$ | Recovery rate from HPV for singly infected individuals | 0.90 | [26] |
| $\rho_L$ | Recovery rate from Chlamydia trachomatis for singly infected individuals | 0.90 | [26] |
| $\rho_{P1}, \rho_{P2}$ | Recovery rate from HPV for dually infected individuals | 0.90 | [26] |
| $\rho_L$ | Recovery rate from HPV for singly infected individuals | 0.90 | assumed |
| $\rho_{L1}, \rho_{L2}$ | Recovery rate from Chlamydia trachomatis for dually infected individuals | 0.90 | assumed |

### 2.1 Basic properties of the model

#### 2.1.1 Positivity and boundedness

For the model (3) to be epidemiologically meaningful, it is important to prove that all its state variables are non-negative for all time (*t*). However, it is to be noted that, since the model (3) monitors the human population, all the parameters of the model are assumed non-negative. The following result can be established:

**Theorem 2.1** *Let the initial data S*_H_ > 0*, I*_HP_ > 0*, I*_SHP_ > 0*, R*_HP_ > 0*, I*_CL_ > 0*, R*_CL_ > 0*, I*_HPCL_ > 0*, I*_SHPCL_ > 0*. Then the solutions* (*S*_H_*, I*_HP_*, I*_SHP_*, R*_HP_*, I*_CL_*, R*_CL_*, I*_HPCL_*, I*_SHPCL_) *of the model* (3) *are positive for all time t >* 0.

*Proof*. Let *t*_1_ = sup*{t >* 0 : *S*_H_ > 0*, I*_HP_ > 0*, I*_SHP_ > 0*, R*_HP_ > 0*, I*_CL_ > 0*, R*_CL_ > 0*, I*_HPCL_ > 0*, I*_SHPCL_ > 0 *∈* [0, *t*]}. Thus, *t*_1_ > 0.

From the first equation of the system (3),

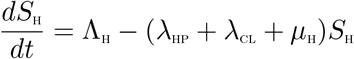

which can be re-written as

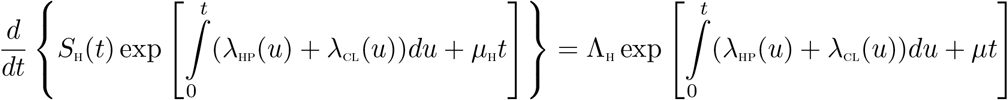

so that

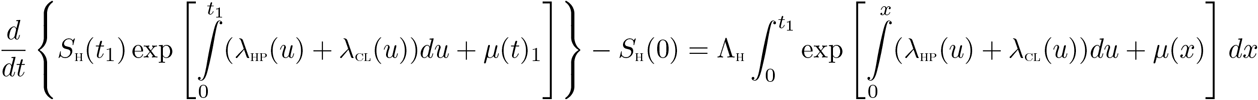

so that

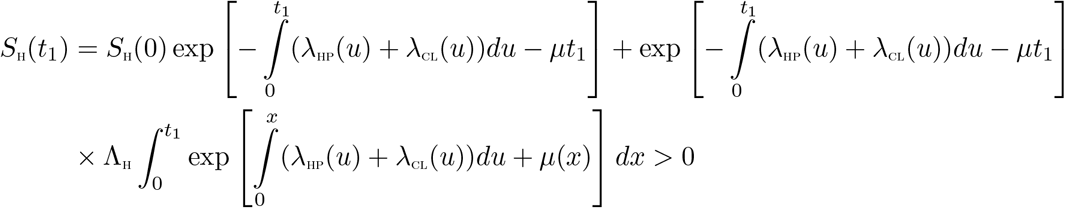

In a similar manner, it can be proven that:

*I*_HP_ > 0*, I*_SHP_ > 0*, R*_HP_ > 0*, I*_CL_ > 0*, R*_CL_ > 0*, I*_HPCL_ > 0*, I*_SHPCL_ > 0

### 2.2 Invariant regions

The co-infection model (3) will be analyzed in a biologically feasible region as follows. Firstly, it is shown that the system (3) is dissipative in a proper subset 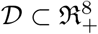. Let

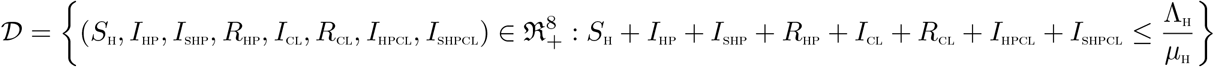

Adding all the equations of the system (3) gives

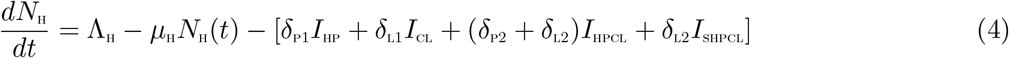

From (4),

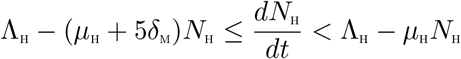

where *δ*_m_ = min*{δ*_P1_*, δ*_P2_*, δ*_L1_*, δ*_L2_*}*

Applying the Comparison theorem [14], it follows that, 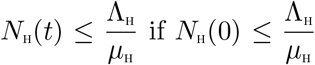. Thus, the region 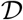 is positively invariant. Hence, it is sufficient to consider the dynamics of the flow generated by the system (3) in 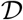. Thus, within this region, the model (3) is said to be epidemiologically and mathematically well-posed [11]. Thus, every solution of the model (3) with initial conditions in 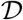 remains in 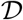 for all time *t* ≥ 0. Therefore, the *ω−*limit sets of the system (3) are contained in 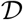. This result is summarized below.

**Lemma 2.1** *The region* 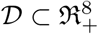 *is positively-invariant for the model* (3) *with initial conditions in* 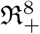.

## 3 Mathematical analysis of the model without controls

In this section, the dynamical properties of the model (3) without controls, are explored. The HPV-only sub-model as well as the full co-infection model shall be considered.

### 3.1 HPV-only Sub-model

Due to complexity of the full co-infection model, certain rigorous analysis, which may not be mathematically feasible for the complete model, are hereby carried out for the HPV-only sub-model. The HPV-only sub-model is obtained from the full co-infection model (3) by setting *I*_CL_ = *R*_CL_ = *I*_HPCL_ = *I*_SHPCL_ = 0. Thus, it is given by:

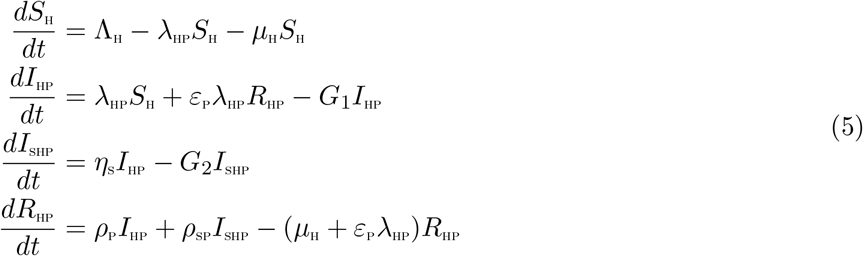

where now,

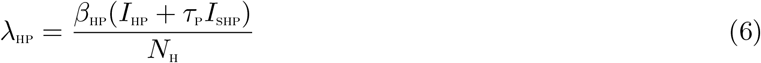

with

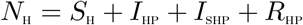

#### 3.1.1 Basic reproduction number of the HPV-only sub-model

The HPV-only sub-model (5) has a DFE, obtained by setting the disease components (*I*_HP_ and *I*_SHP_) as well as the right-hand sides of the equations in the model (5) to zero, given by

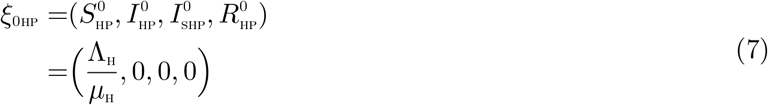

The basic reproduction number, using the next generation operator method [45], is given by

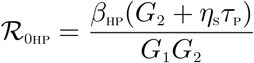

with

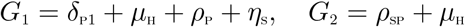

#### 3.1.2 Existence of Endemic Equilibrium of the HPV-only sub-model

In this section, the existence of an endemic equilibrium of the HPV-only sub-model shall be investigated, since this can not be shown for the full co-infection model (due to complexity).

Let an arbitrary equilibrium point of the HPV-only sub-model be represented by

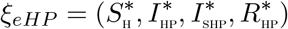

The steady state solutions of equations of the sub-model (5) are given by:

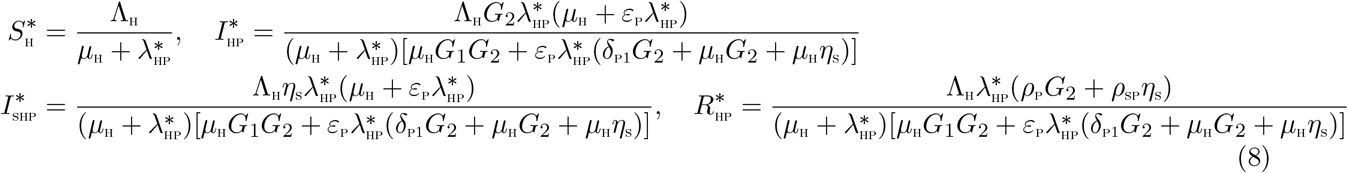

Substituting the above expressions into the force of infection (6), at steady state, gives the following polynomial:

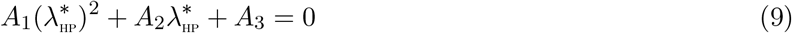

with,

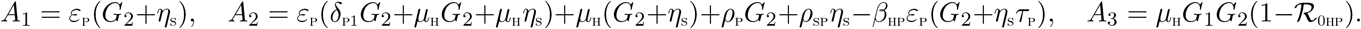

It is observed from (9), that the coefficient *A*_1_, is always positive and *A*_3_ is positive (negative) if 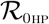 is less (greater) than unity. Hence, the following result can be established:

**Theorem 3.1** *The sub-model model* (5) *has*

i. *a unique endemic equilibrium if* 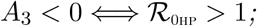
ii. *a unique endemic equilibrium if A*_2_ < 0 *and A*_1_ = 0 *or* 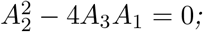
iii. *two endemic equilibria if A*_1_ > 0*, A*_2_ < 0 *and* 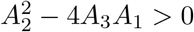 and 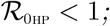;
iv. *no endemic equilibrium otherwise*.

The third item (iii) of the above theorem suggests the possibility of a backward bifurcation in the HPV-only sub-model. The associated backward bifurcation diagram is presented in Figure 2. It is imperative to note that, setting the HPV re-infection term *ε*_P_ = 0, reduces the quadratic (9) to 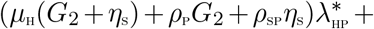 *A*_3_ = 0, resulting in no sign changes in the polynomial equation (9), as (*μ*_H_(*G*_2_ + *η*_s_) + *ρ*_P_*G*_2_ + *ρ*_SP_*η*_s_) > 0 and *A*_3_ > 0 (for 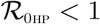). Hence no existence of an endemic equilibrium for 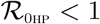, ruling out the existence of backward bifurcation in the HPV-only sub-model (5) in the absence of HPV re-infection.

**Figure 2:**
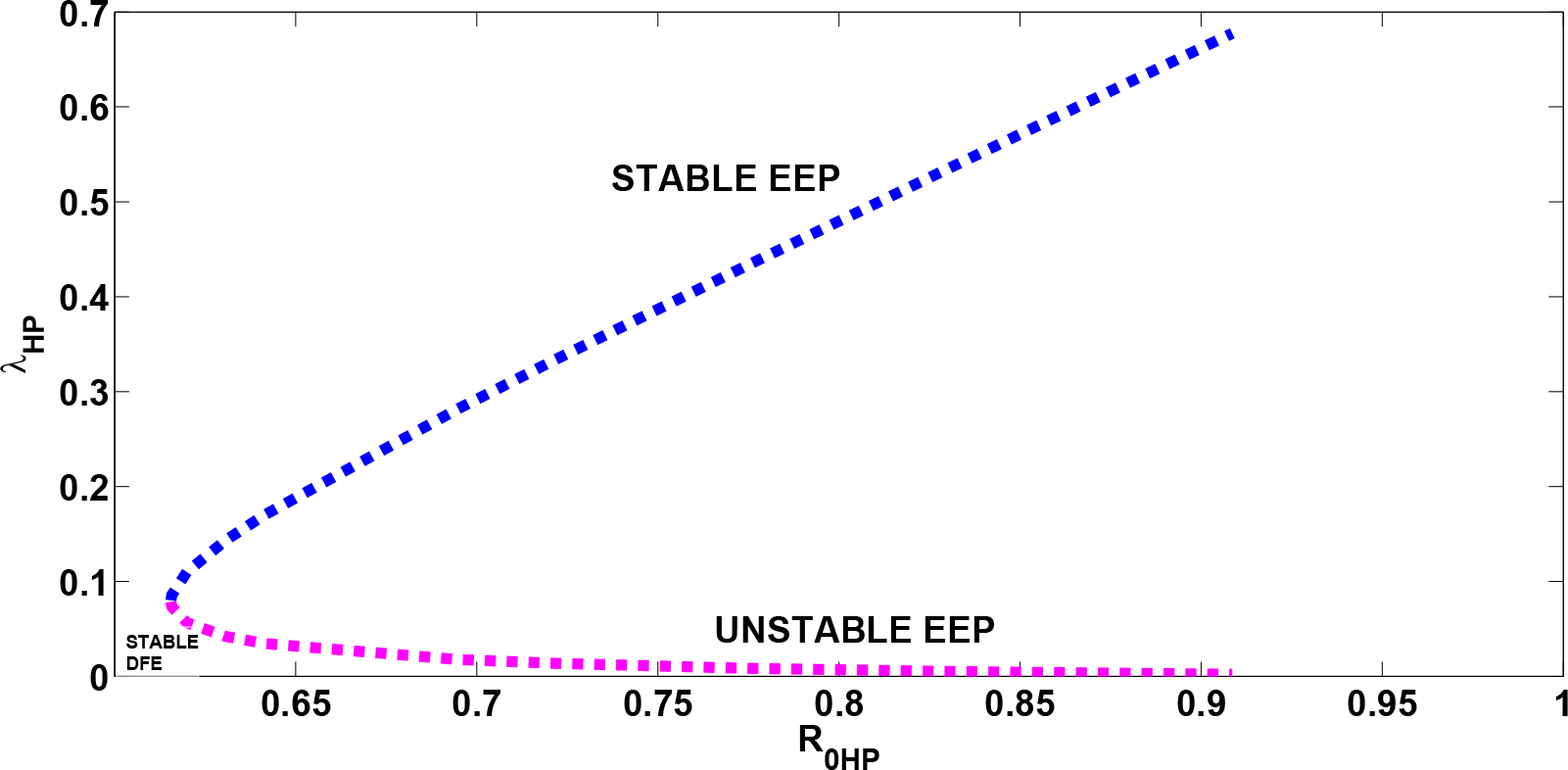
Bifurcation diagram for the HPV-only sub-model (5). Parameter values used are: 1.15168 ≤ *β*_HP_ *≤* 1.70*, ε*_P_ = 1.0. All other parameters as in Table 2

### 3.2 Analysis of the full co-infection model

In this section, the qualitative properties of the full co-infection model (3) without controls, is studied.

### 3.3 Basic reproduction number of the full co-infection model (3)

The HPV-Chlamydia trachomatis co-infection model (3) has a DFE, obtained by setting the right-hand sides of the equations in the model (3) as well as the disease classes (*I*_HP_*, I*_SHP_*, I*_CL_*, I*_HPCL_*, I*_SHPCL_) to zero, given by

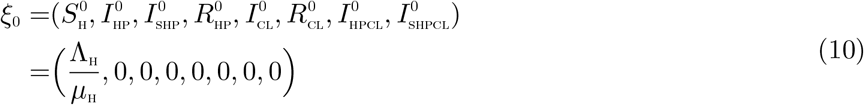

The basic reproduction number of the HPV-Chlamydia trachomatis co-infection model (3), using the approach illustrated in [45], is given by 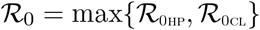 where 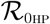 and 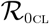 are, respectively, the HPV and Chlamydia trachomatis associated reproduction numbers, given by

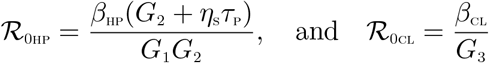

where

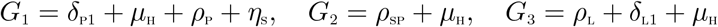

### 3.4 Local asymptotic stability of disease-free equilibrium (DFE) of the co-infection model (3)

**Lemma 3.1** *The DFE, ξ*_0_*, of the HPV-Chlamydia trachomatis co-infection model* (3) *is locally asymptotically stable if* 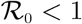, *and unstable if* 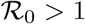.

**Proof**

The local stability of the HPV-Chlamydia co-infection model is analysed by the Jacobian matrix of the system (3) at *ξ*_0_, given by:

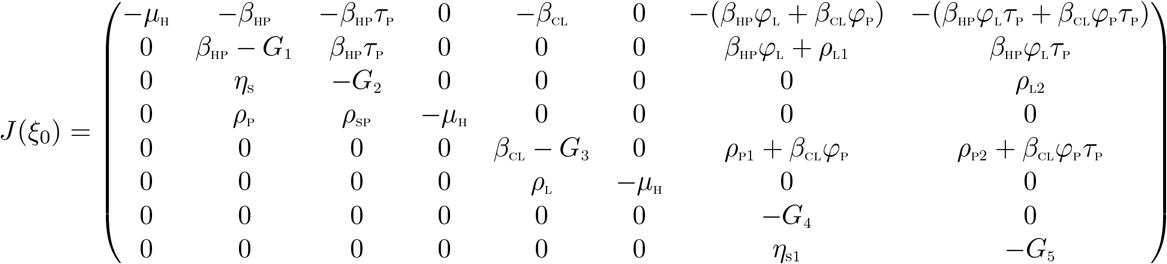

where,

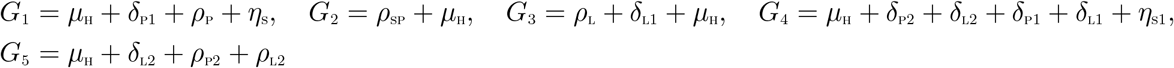

The eigenvalues are *λ*_1_ = *−μ*_H_, *λ*_1_ = *−μ*_H_, *λ*_3_ = *−μ*_H_, *λ*_4_ = *−*(*μ*_H_+*δ*_P2_+*δ*_L2_+*ρ*_L1_+*η*_S1_), *λ*_5_ = *−*(*μ*_H_+*δ*_L2_+*ρ*_P2_+*ρ*_L2_), 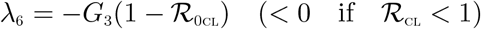 and the solutions of the characteristic polynomial:

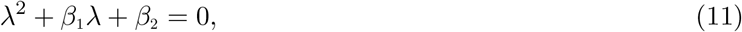

where

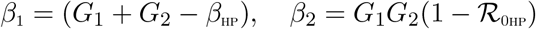

Applying the Routh-Hurwitz criterion, the quadratic equation (11) will have roots with negative real parts if and only if *β*_1_ > 0, *β*_2_ > 0 and *β*_1_*β*_2_ > 0. It can be shown that, 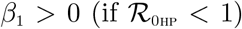. This is true, as *β*_HP_(*G*_2_ + *η*_S_*τ*_P_) *< G*_1_*G*_2_ = *⇒ β*_HP_ *< G*_1_. Thus it follows, that *G*_1_ + *G*_2_ > *β*_HP_ (since the model parameters are assumed non-negative). Also, *β*_2_ > 0 (if 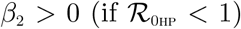). Moreover, *β*_1_*β*_2_ > 0. As a result, the disease-free equilibrium, *ξ*_0_ is locally asymptotically stable if 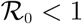.

### 3.5 Global asymptotic stability(GAS) of the disease-free equilibrium(DFE) *ξ*_0_ of the co-infection model

The approach illustrated in [4] is used to investigate the global asymptotic stability of the disease free equilibrium of the co-infection model. In this section, two conditions are listed, that if met, they guarantee the global asymptotic stability (GAS) of the disease-free equilibrium (DFE). Firstly, system (3) must be written in the form:

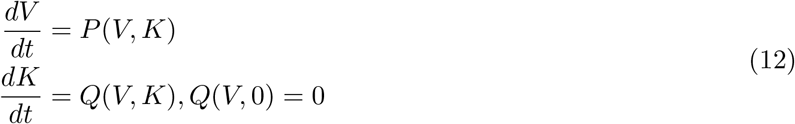

where *V* ∈ *R^m^* denotes (its components) the number of uninfected individuals and *K* ∈ *R^n^* denotes (its components) the number of infected individuals. *U*_0_ = (*V^∗^*, 0) denotes the disease-free equilibrium of this system. The conditions (*W* 1) and (*W* 2) below must be satisfied in order to guarantee local asymptotic stability:

(*W* 1): For 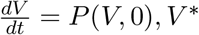 is globally asymptotically stable (GAS),
(*W* 2): 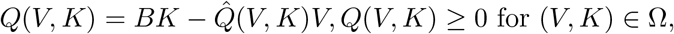

where *B* = *D_K_Q*(*V^∗^*, 0) is an M-matrix (the off-diagonal elements of *B* are nonnegative) and Ω is the region where the model makes biological sense. If system (3) satisfies the above two conditions then the following theorem holds:

**Theorem 3.2** *The fixed point U*_0_ = (*V^∗^*, 0) *is a globally asymptotic stable (GAS) equilibrium of* (3) *provided that R*_0_ < 1 *(LAS) and that assumptions* (*W* 1) *and* (*W* 2) *are met*

**Proof**

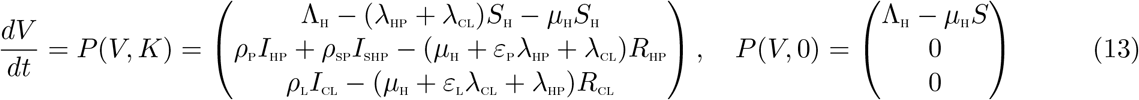

where *V* denotes the number of non-infectious compartments and *K* denotes the number of infectious compartments

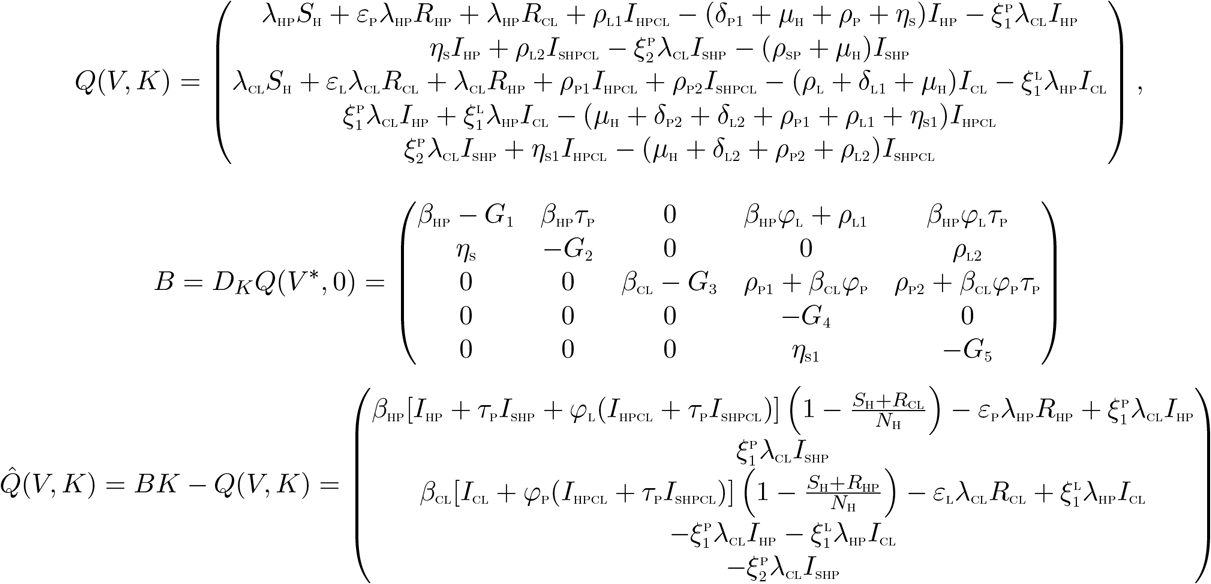

It is clear from the above, that, 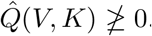. Hence the DFE may not be globally asymptotically stable, suggesting the possibility of a backward bifurcation. This supports the backward bifurcation analysis in the proceeding section.

### 3.6 Backward bifurcation analysis of the co-infection model (3)

In this section, the type of bifurcation the model (3) will exhibit is determined, using the approach illustrated by Castillo-Chavez and Song [5]. Backward bifurcation analysis also has been carried out in several disease models [8, 22, 24, 27, 36, 37]. The result below is established.

**Theorem 3.3** *Suppose a backward bifurcation coefficient a >* 0*, (with a defined below), when* 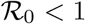

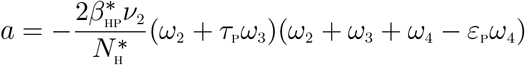

*then model* (3) *undergoes the phenomenon of backward bifurcation at* 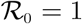. *If a* < 0*, then the system* (3) *exhibits a forward bifurcation at* 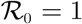.

**Proof**

Suppose

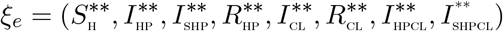

represents any arbitrary endemic equilibrium of the model. The existence of backward bifurcation will be studied using the Centre Manifold Theory [5]. To apply this theory, it is appropriate to make the following change of variables.

Let

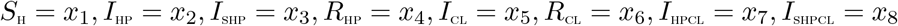

Moreover, using the vector notation

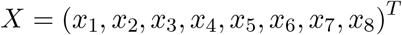

the model (3) can be re-written in the form

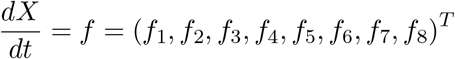

as follows:

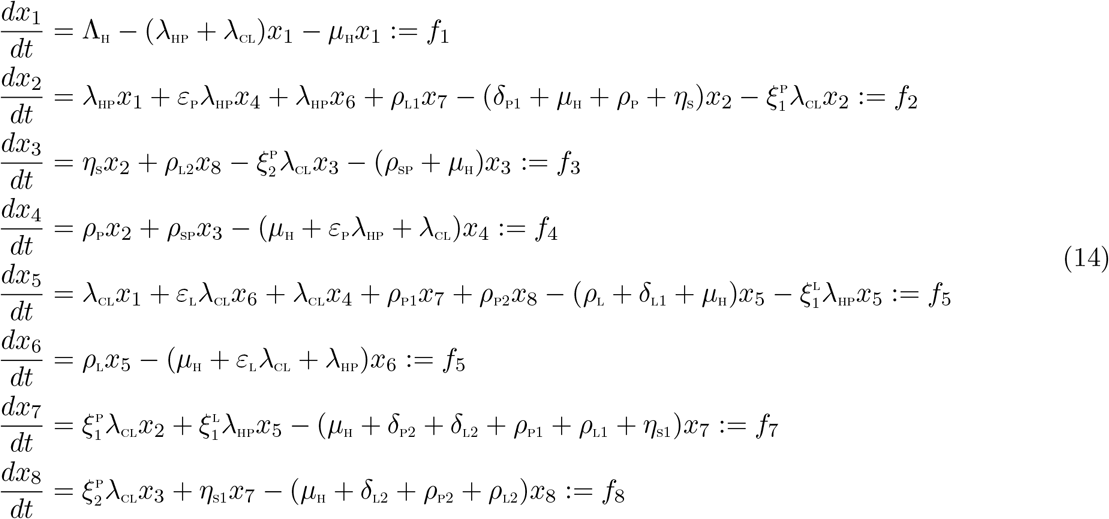

with

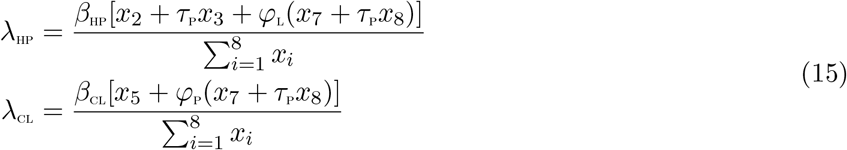

Consider the case when 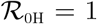. Assume, further, that *β*_HP_ is chosen as a bifurcation parameter. Solving for 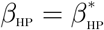 from 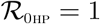 gives

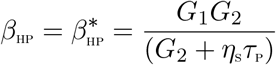

Evaluating the Jacobian of the system (14) at the DFE, *J*(*ξ*_0_), and evaluating the right eigenvector, **w** = [*ω*_1_*, ω*_2_*, ω*_3_*, ω*_4_*, ω*_5_*, ω*_6_*, ω*_7_*, ω*_8_]^*T*^, associated with the simple zero eigenvalue of *J*(*ξ*_0_), gives

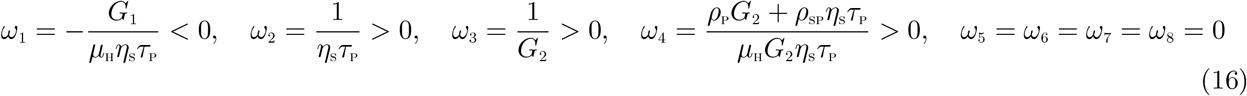

Likewise, the components of the left eigenvector of 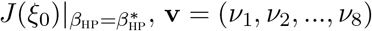, satisfying **v.w** = 1 are

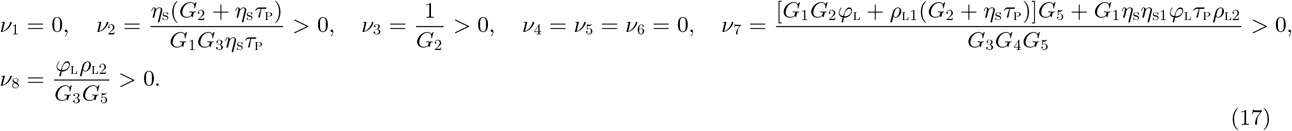

The non-zero second partial derivatives of the functions *f_i_*(*i* = 1*, …*, 8) are given by

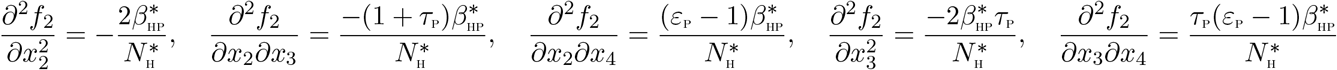

The associated bifurcation coefficients defined by *a* and *b*, are given by:

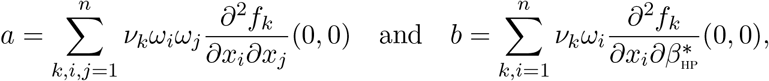

are computed to be

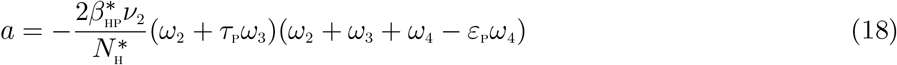

and

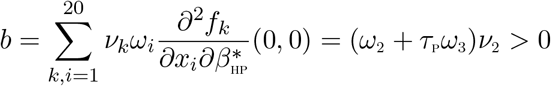

Since the bifurcation coefficient *b* is positive, it follows from Theorem 4.1 in [5] that the model (3), or the transformed model (14), will undergo the phenomenon of backward bifurcation if the coefficient, *a*, given by (18) is positive. Setting the HPV re-infection term *ε*_P_ = 0, it is well observed that the bifurcation coefficient, *a* < 0. Hence, backward bifurcation does not occur in the HPV-Chlamydia co-infection model, in the absence of HPV re-infection. The epidemiological interpretation is that if recovery from HPV does not confer lifelong immunity, then the control of HPV-Chlamydia trachomatis becomes difficult, even when the associated reproduction number 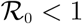.

## 4 Analysis of the optimal control model

In this section, the Pontryagin’s Maximum Principle is used to determine the necessary conditions for the optimal control of the oncogenic HPV-Chlamydia co-infection model. Time dependent controls are incorporated into the model (3) to determine the optimal strategy for curbing the co-infections of the two diseases. Thus,

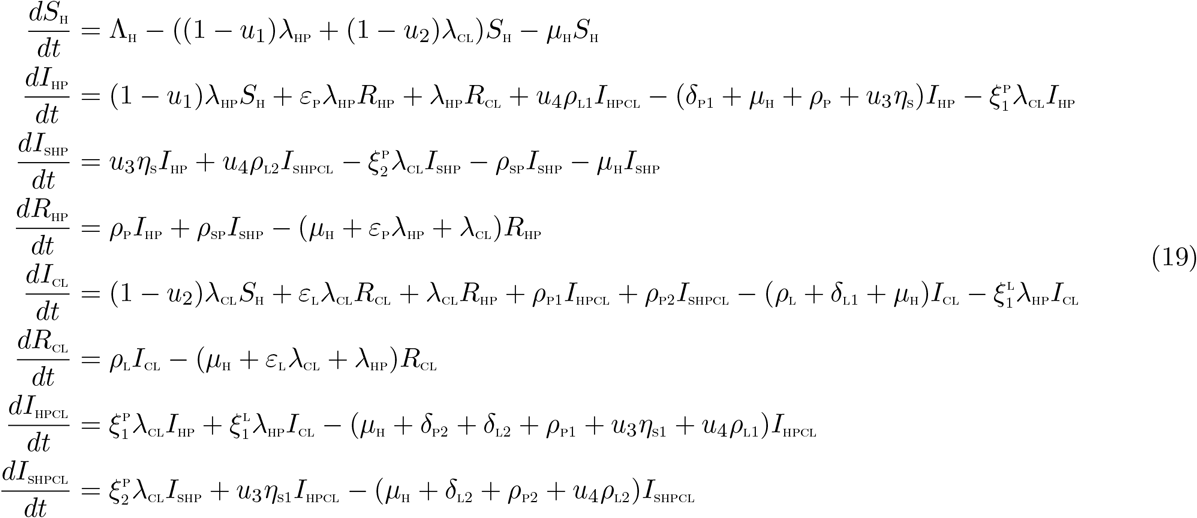

subject to the initial conditions 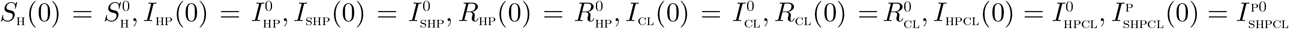
with:

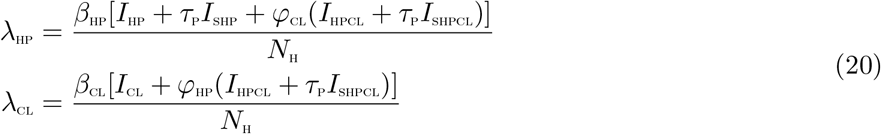

The control functions, *u*_1_(*t*)*, u*_2_(*t*)*, u*_3_(*t*) and *u*_4_(*t*) are bounded, Lebesgue integrable functions. The control *u*_1_(*t*) and *u*_2_(*t*) represent the efforts (such as HPV vaccination, sexual abstinence, monogamous relationship with an uninfected partner and condom use by sexually active susceptible individuals) aimed at preventing incident HPV and Chlamydia infections, respectively. The control *u*_3_(*t*) is the effort aimed at screening of HPV infected individuals so as to reduce their transmission probability. Chlamydia treatment control for individuals dually infected with HPV and Chlamydia is denoted by *u*_4_(*t*). The controls *u*_1_ and *u*_2_ satisfies 0 ≤ *u*_1_*, u*_2_ *≤* 0.9, the control *u*_3_ satisfies 0 < *u*_3_ ≤ 1, whereas the control *u*_4_ satisfies 0 < *u*_4_ ≤ *θ*, where *θ* is the Chlamydia drug efficacy used for the treatment of co-infected individuals. Our optimal control problem involves a situation where the number of HPV-infected, Chlamydia-infected, the co-infection cases and the cost of implementing preventive, screening and treatment controls *u*_1_(*t*)*, u*_2_(*t*)*, u*_3_(*t*) and *u*_4_(*t*) are minimized subject to the state system (19). For this, the objective functional below, is considered.

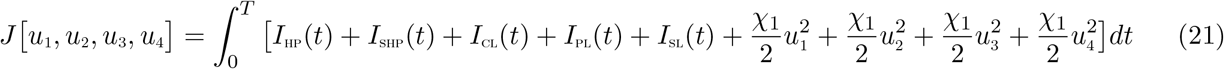

*T* is the final time. An optimal control, 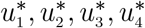, is to be found, such that

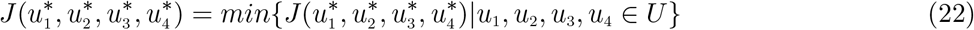

where 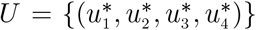 such that 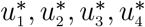 are measurable with 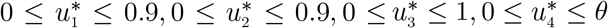 for *t* ∈ [0, *T*] is the control set.

### 4.1 Existence of Optimal Control

The existence of such an optimal solution which minimizes the objective functional *J* is now established.

**Theorem 4.1** *Given the objective functional J, defined on the control set U, and subject to the state system* (19) *with non-negative initial conditions at t* = 0*, then there exists an optimal control triple u^∗^* = (*u*_1_*, u*_2_*, u*_3_*, u*_4_) *such that J*(*u^∗^*) = *min {J*(*u*_1_*, u*_2_*, u*_3_*, u*_4_)*|u*_1_*, u*_2_*, u*_3_*, u*_4_ *∈ U}*.

Let *U* = [0, 1]^4^ be the control set, *υ* = (*u*_1_*, u*_2_*, u*_3_*, u*_4_) *∈ U*, *x* = (*S*_H_*, I*_HP_*, I*_SHP_*, R*_HP_*, I*_CL_*, R*_CL_*, I*_HPCL_*, I*_SHPCL_) and *f*(*t, x, υ*) be the right hand of (19), that is

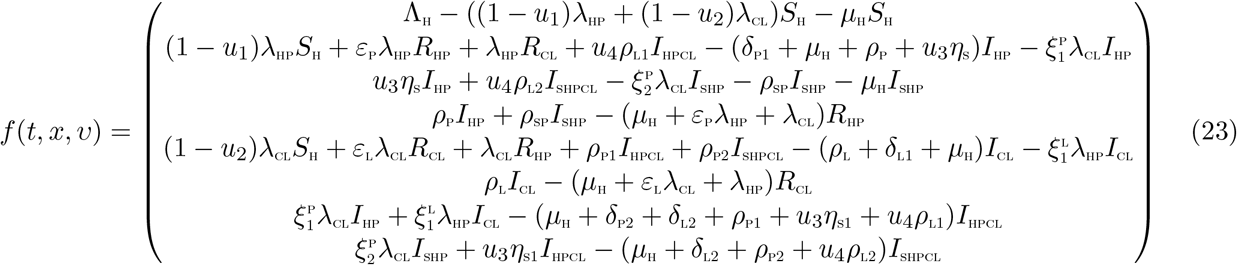

To prove Theorem 4.1, it is necessary to verify the following conditions proposed by Fleming and Rishel [9]:

i. The solution set for the model system (19) with corresponding control functions in *U* is non empty: To establish the existence of a solution corresponding to every admissible control in *U*, it is required to show that the state variables associated with the state equations are bounded and the state equations are continuous and Lipschitz in state variables. Clearly, it is observed that all the state equations are continuous in state variables. Moreover, since the total population *N*_H_(*t*) is bounded above by 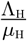, it follows that the state variables are bounded above by 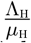. Equally, the Lipschitz condition with respect to state variables follows from the boundedness of the partial derivatives with respect to state variables in the state system. Consequently, the set of all solutions of the control system (19) is non-empty.
ii. The control model system can be expressed as a linear function of control variables (*u*_1_*, u*_2_*, u*_3_*, u*_4_), with the coefficients as functions of time and state variables:

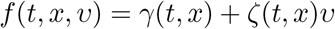

with

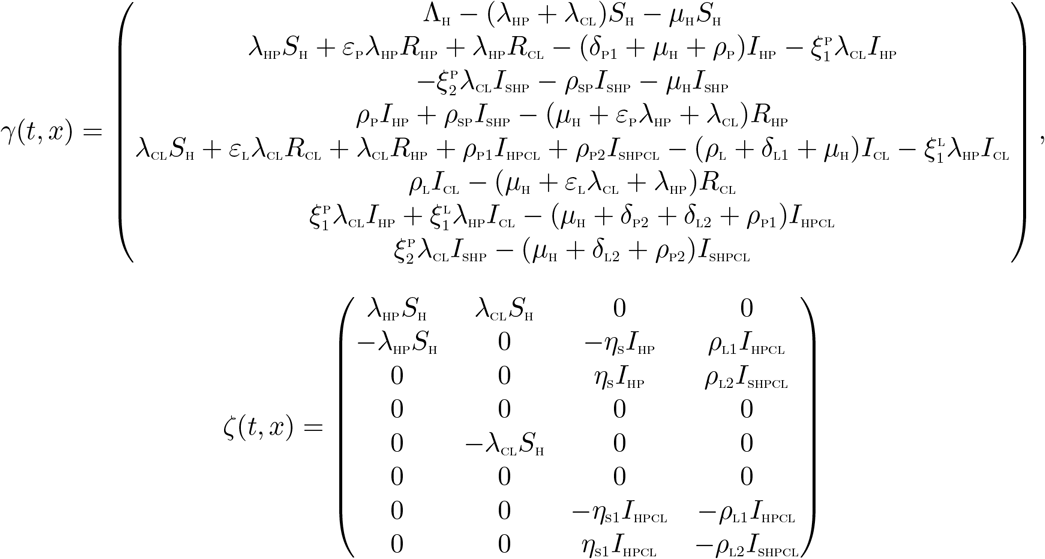
iii. There exists constants *α*_1_*, α*_2_ and *α*_3_ such that the Lagrangian of the problem, 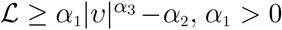, *α*_2_ > 0, *α*_3_ > 1

The Lagrangian of the problem (19) is given as 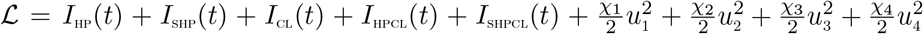. The lagrangian, 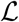, is a quadratic function of *υ* = (*u*_1_*, u*_2_*, u*_3_*, u*_4_) and hence convex on *U*. The bound on 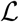 is now established. Note that 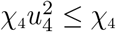 since *u*_4_ *∈* [0, 1], so that 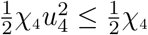. Now,

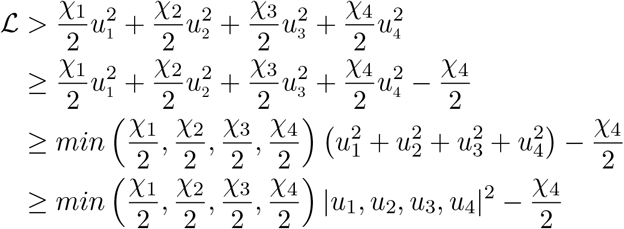

Hence,

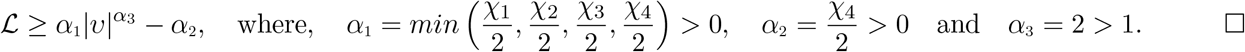

The Pontryagin’s Maximum Principle [30] gives the necessary conditions which an optimal control pair must satisfy. This principle transforms (19), (21) and (22) into a problem of minimizing a Hamiltonian, 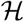, pointwisely with regards to the control functions, *u*_1_*, u*_2_*, u*_3_*, u*_4_:

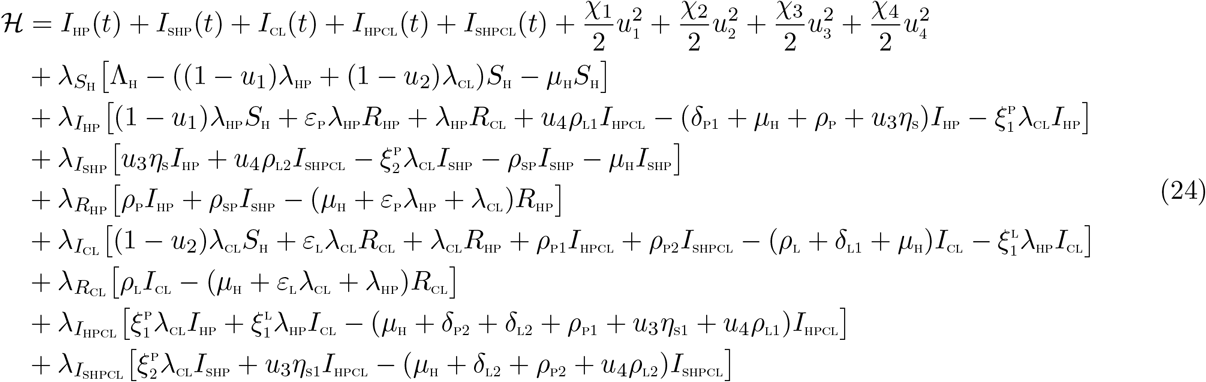

**Theorem 4.2** *For an optimal control set u*_1_*, u*_2_*, u*_3_*, u*_4_ *that minimizes J over U, there are adjoint variables*, *λ*_1_*, λ*_2_*, …, λ*_8_ *satisfying*

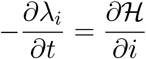

*and with transversality conditions*

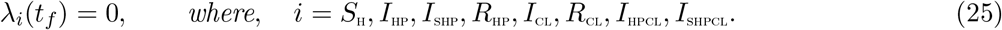

*Furthermore*,

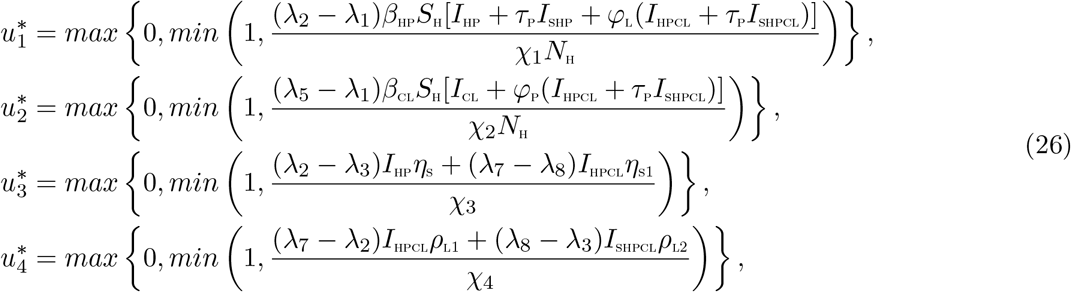

**Proof of Theorem 4.2**

Suppose 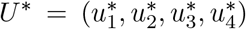 is an optimal control and 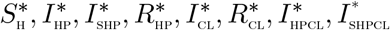 are the corresponding state solutions. Applying the Pontryagin’s Maximum Principle [30], there exist adjoint variables satisfying:

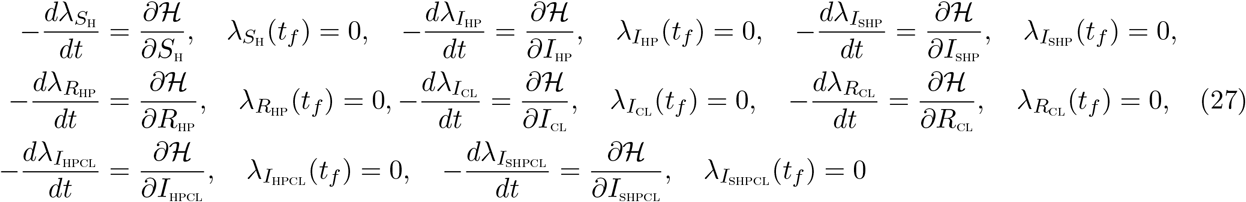

with transversality conditions;
*λ_S_*_H_(*t_f_*) = *λ_I_*_HP_(*t_f_*) = *λ_I_*_SHP_(*t_f_*) = *λ_R_*_HP_(*t_f_*) = *λ_I_*_CL_(*t_f_*) = *λ_R_*_CL_(*t_f_*) = *λ_I_*_HPCL_(*t_f_*) = *λ_I_*_SHPCL_(*t_f_*) = 0 The behaviour of the control can be determined by differentiating the Hamiltonian, 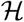 with respect to the controls(*u*_1_*, u*_2_*, u*_3_*, u*_4_) at *t*. On the interior of the control set, where 0 *< u_j_* < 1 for all (*j* = 1, 2, 3, 4),

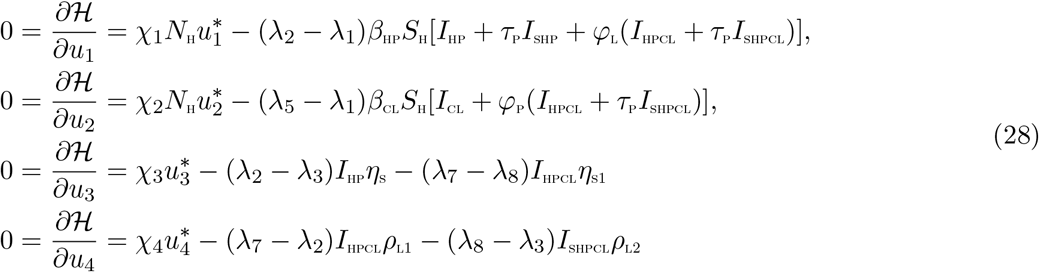

Therefore, the following is obtained [15]

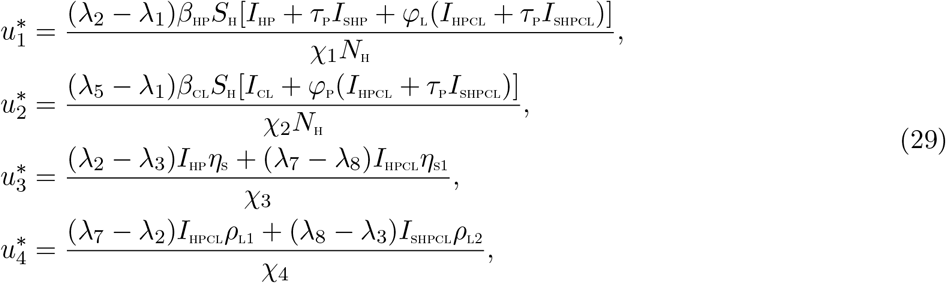

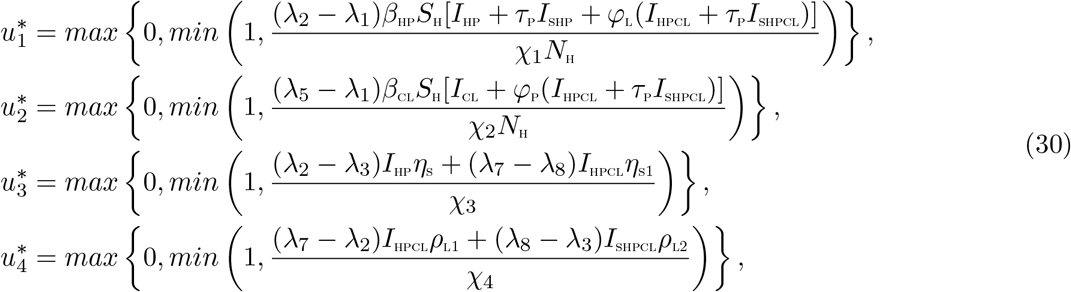

## 5 Simulations

In this section, uncertainty and sensitivity analyses of the parameters of the model are carried out due to imprecision which may arise from the estimates of some of the parameters in the model. It is imperative to state here that, very limited data is available on the co-infection of HPV and Chlamydia trachomatis. Numerical simulations be carried out on the optimal control model (19), in order to assess the effect of different interventions on the dynamics of the co-infections of HPV and Chlamydia trachomatis.

### 5.1 Uncertainty and sensitivity analyses

As a result of the uncertainties which are expected to come up in parameter estimates used in the numerical simulations, a Latin Hypercube Sampling (LHS) [2] is implemented on the parameters of the model. For the sensitivity analysis, a Partial Rank Correlation Coefficient (PRCC) was carried out. 1,000 simulations of the co-infection model (3) *per* LHS were run. Using the HPV associated basic reproduction number, 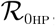, as the response function, it is observed in Table 1 that the three top-ranked parameters that drive the dynamics of the co-infection model are effective contact rate for HPV transmission, *β*_HP_ and the recovery rate from HPV, *ρ*_P_, and the modification parameter accounting for the infectiousness of individuals who have undergone HPV screening, *τ*_P_. In addition, using the Chlamydia trachomatis associated reproduction number, 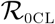 as the response function, the two key parameters that drive the dynamics of the model are the effective contact rate for Chlamydia trachomatis transmission, *β*_CL_ as well as the recovery rate from Chlamydia trachomatis *ρ*_L_.

Using the total number of individuals infected with HPV (*I*_HP_) as the response function, the parameters that strongly drive the dynamics of the HPV-Chlamydia trachomatis co-infection model (3) are the effective contact rate for HPV transmission, *β*_HP_ and the recovery rate from Chlamydia trachomatis infection for dually infected individuals, *ρ*_L2_. When total population of individuals infected with Chlamydia trachomatis (*I*_CL_) is used as the response function, the parameters that strongly drive the dynamics of the HPV-Chlamydia trachomatis co-infection model (3) are the effective contact rate for HPV transmission, the effective contact rate for Chlamydia trachomatis transmission, *β*_CL_, the recovery rate from HPV infection for dually infected individuals, *ρ*_P1_. Finally, using the population of individuals dually infected with HPV and Chlamydia trachomatis (*I*_HPCL_) as the input, the eight highly ranked parameters that influence the dynamics of the co-infection model are the effective contact rate for HPV transmissibility, *β*_HP_, the effective contact rate neccesary for Chlamydia trachomatis transmission, *β*_CL_, HPV screening rate for dually infected individuals, *η*_S1_, the modification parameters accounting for increased infectiousness of dually infected individuals, *φ*_P_ and *φ*_L_, respectively, the modification parameter accounting for the infectiousness of individuals who have undergone HPV screening, *τ*_P_, and the modification parameters accounting for increased susceptibility to HPV and Chlamydia trachomatis infections, 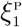 and 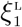 respectively.

### 5.2 Numerical simulations

Numerical simulations of the optimal control problem (19), adjoint equations (27) and characterizations of the control (30) are implemented by the Runge Kutta method using the forward backward sweep (carried out in MATLAB). The algorithm used for the solution of the state system (19) is based on the approach proposed in [15]. The weight constants are assumed to be: *χ*_1_ = 500*, χ*_2_ = 500*, χ*_3_ = 400 and *χ*_4_ = 400. Demographic data relevant to the dynamics of the co-infection of HPV and Chlamydia in Uganda are used [42]. The initial conditions are assumed to be: *S*_H_(0) = 10000*, I*_HP_(0) = 2000*, I*_SHP_(0) = 2500*, R*_HP_(0) = 2000*, I*_CL_(0) = 2000*, R*_CL_(0) = 2000*, I*_HPCL_(0) = 2500*, I*_SHPCL_(0) = 2000. Based on the sensitivty analysis results in Section 5.1, the following four different control strategies are implemented for the numerical simulations of the co-infection model (19).

i. Strategy A: HPV prevention (*u*_1_ ≠ 0) and Chlamydia trachomatis prevention (*u*_2_ ≠ 0);
ii. Strategy B: HPV prevention (*u*_1_ ≠ 0) and screening of HPV infected individuals (*u*_3_ ≠ 0);
iii. Strategy C: Chlamydia trachomatis prevention (*u*_2_ ≠ 0) and treatment (*u*_4_ ≠ 0).
iv. Strategy D: HPV screening (*u*_3_ ≠ 0) and Chlamydia trachomatis treatment (*u*_4_ ≠ 0).

#### 5.2.1 Strategy A: HPV prevention (*u*_1_ ≠ 0) and Chlamydia trachomatis prevention (*u*_1_ ≠ 0) controls

Simulations of the optimal control system (19) when HPV prevention (*u*_1_ ≠ 0) and Chlamydia trachomatis prevention (*u*_2_ ≠ 0) controls are applied, are shown in Figure 3. It is observed that when this control strategy is implemented, there is a significant reduction in the total number of individuals singly infected with HPV (Figure 3 (a)), total number of individuals singly infected with Chlamydia trachomatis (Figure 3b) and the total number of individuals dually infected with HPV and Chalmydia trachomatis (Figure 3 (c)). Particularly, in addition to averting 64,133 new cases of HPV infections and preventing 35,660 new cases of Chlamydia trachomatis infection, this control strategy also averts 3,364 new co-infection cases. The control profile for this strategy given in Figure 4 (a) shows that control *u*_1_ is at its peak for the first 4.2 years and ultimately declines to zero at final time (when *t* = 5 years). In a similar manner, the control *u*_2_ is at the upper bound for the first 4.6 years before finally declining to zero. The cost function for the combined effects of these two controls is given by Figure 4 (b).

**Figure 3:**
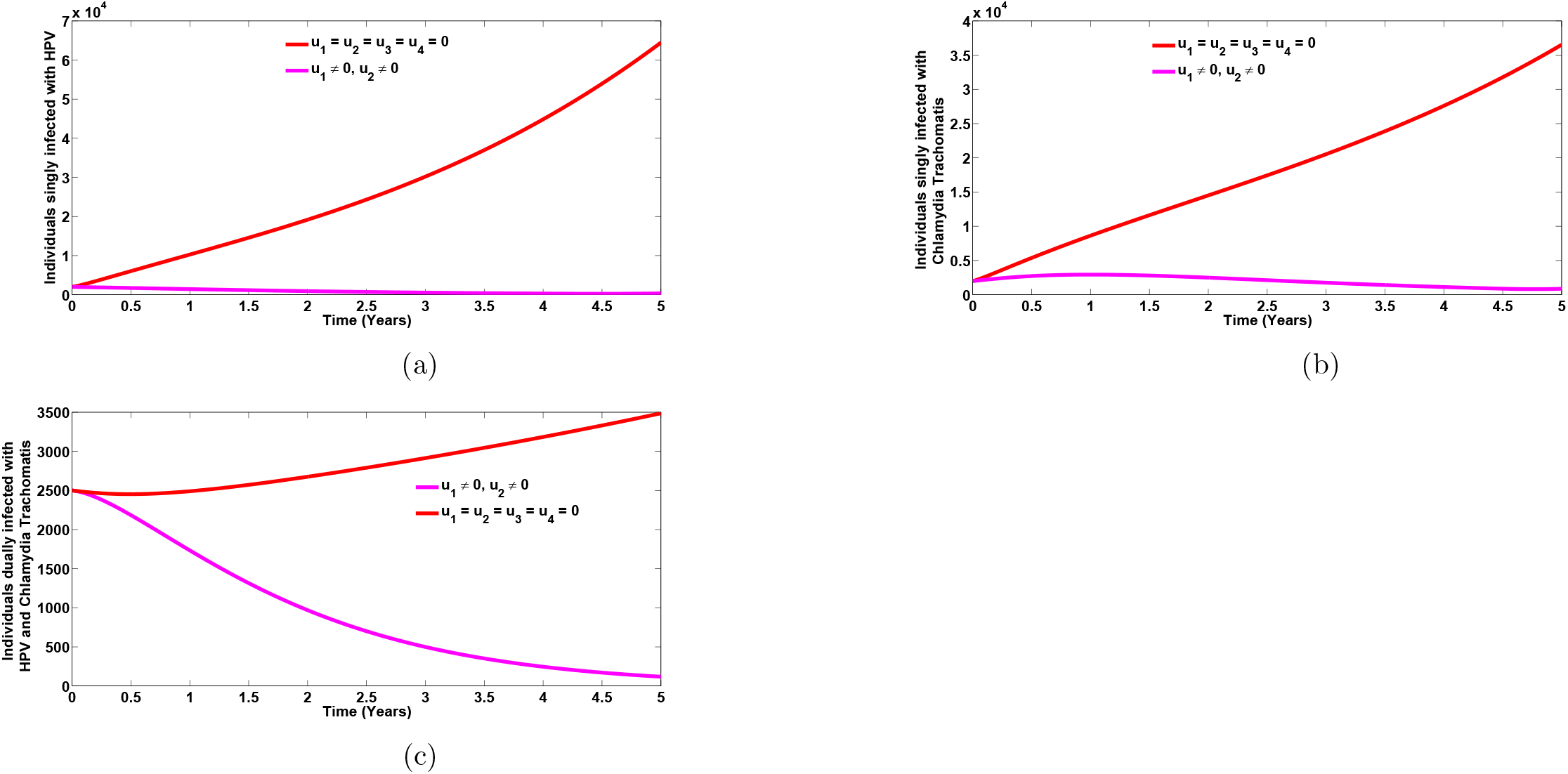
Plots of the total number of individuals singly infected with HPV (Figure 3 (a)), total number of individuals singly infected with Chlamydia trachomatis (Figure 3 (b)), as well as the total number of individuals dually infected with HPV and Chlamydia trachomatis (Figure 3 (c)), in the presence of HPV prevention (*u*_1_ ≠ 0) and Chlamydia trachomatis prevention (*u*_2_ ≠ 0) controls. Here, *β*_HP_ = 1.35*, β*_CL_ = 1.0. All other parameters are as in Table 2

**Figure 4:**
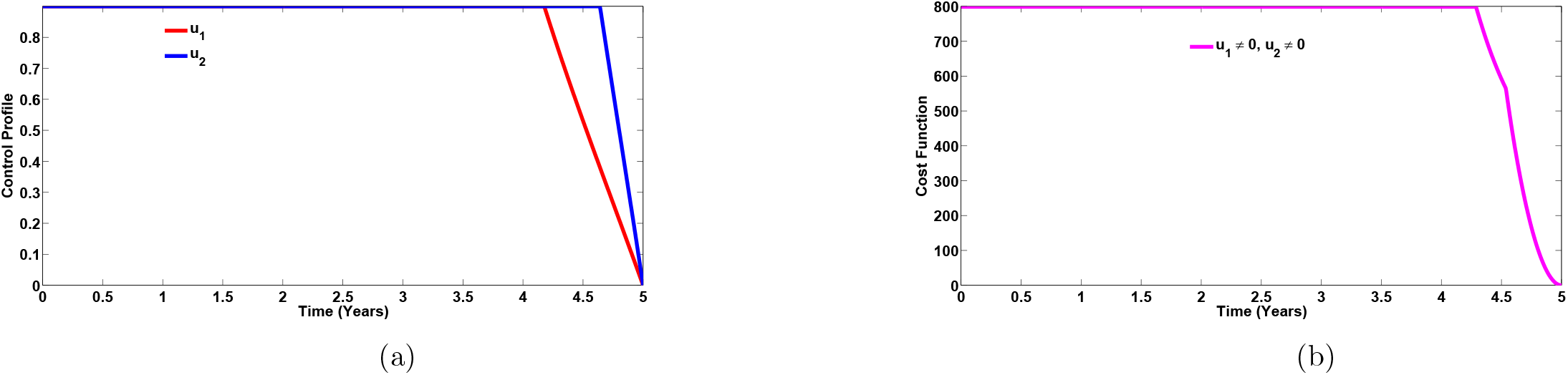
Control profile (Figure 4 (a)) and cost function (Figure 4 (b)) for the combined effects of the controls *u*_1_ and *u*_2_ on the dynamics of the HPV-Chlamydia trachomatis co-infection model (3). Here, *β*_HP_ = 1.35*, β*_CL_ = 1.0. All other parameters are as in Table 2

#### 5.2.2 Strategy B: HPV prevention (*u*_1_ ≠ 0) and screening (*u*_3_≠ 0) controls

The simulations of the total number of infected individuals in the presence of HPV-only intervention controls (prevention (*u*_1_) and screening (*u*_3_)) are depicted in Figures 5(a)–5(c). Applying this intervention strategy, it is observed that the total number of individuals singly infected with HPV (Figure 5 (a)) and the total number of individuals dually infected with HPV and Chlamydia trachomatis (Figure 5 (c)), respectively, decrease significantly in comparison to when no control strategy is applied. However, a positive population level impact is observed on the total number of individuals singly infected with Chlamydia trachomatis (Figure 5(b)). Specifically, in addition to averting 64,132 new HPV cases, this strategy equally prevents 6,570 new cases of Chlamydia trachomatis infection. This intervention strategy also averts 3,347 new coinfection cases. This result conforms with the epidemiological findings reported in Section 1 that higher prevalence of Chlamydia trachomatis infection has been observed in HPV infected individuals [10, 16]. Hence, focusing only on HPV controls, can in turn bring down the burden of the Chlamydia trachomatis as well as the co-infection of the two diseases. The control profile for this strategy presented in Figure 6 (a) shows that control *u*_1_ is at its peak for the first 4.4 years and ultimately declines to zero at time, *t* = 5 years. In a similar manner, the control *u*_3_ is at the maximum level of 100% for the first 0.75 year before gradually declining to zero at time, *t* = 5 years. The cost function for the combined effects of these two controls is given by Figure 6 (b).

**Figure 5:**
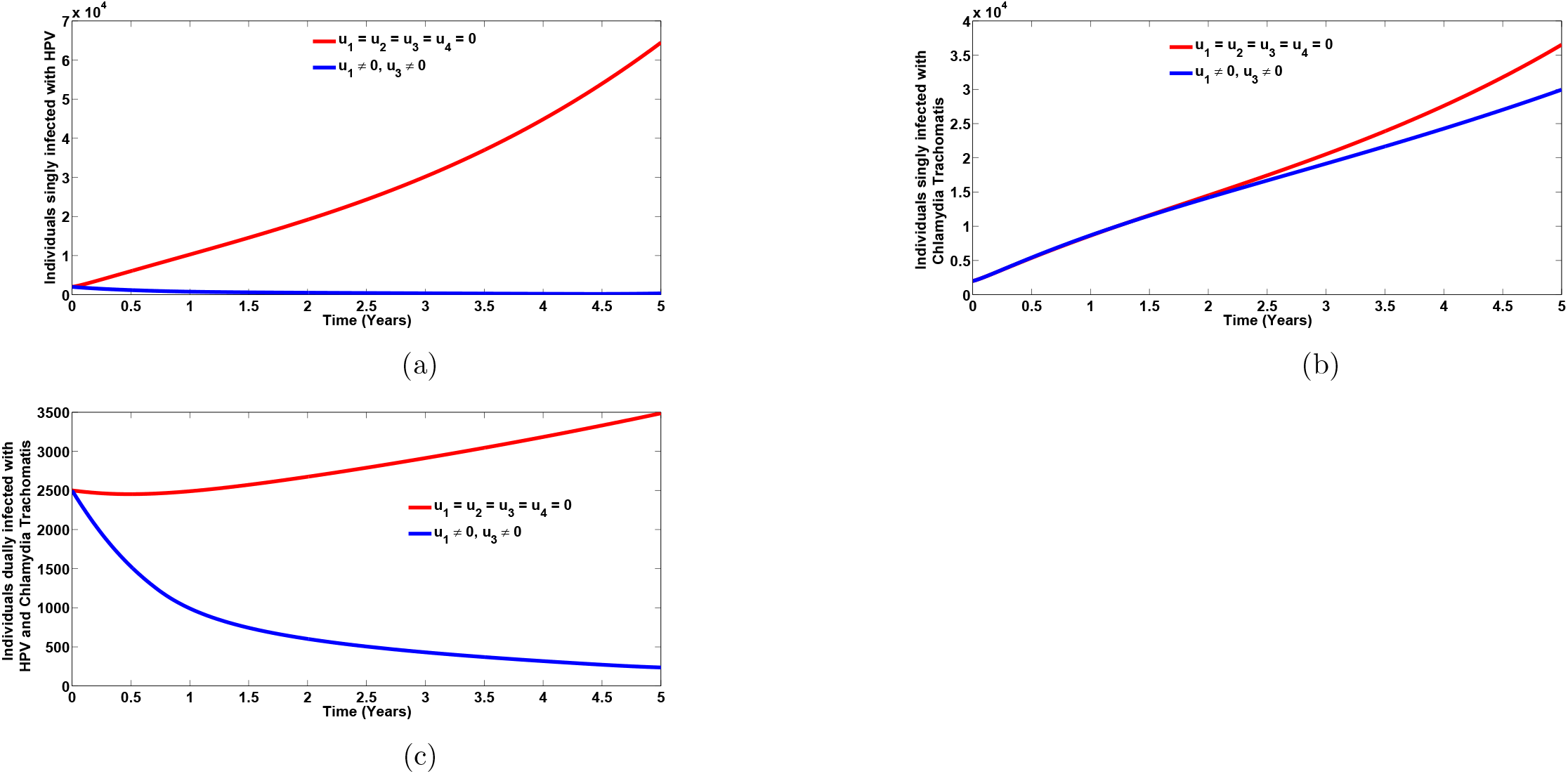
Plots of the total number of individuals singly infected with HPV (Figure 5 (a)), total number of individuals singly infected with Chlamydia trachomatis (Figure 5 (b)), as well as the total number of individuals dually infected with HPV and Chlamydia trachomatis (Figure 5 (c)), in the presence of HPV prevention (*u*_1_ ≠ 0) and screening (*u*_3_ ≠ 0) controls. Here, *β*_HP_ = 1.35*, β*_CL_ = 1.0. All other parameters are as in Table 2

**Figure 6:**
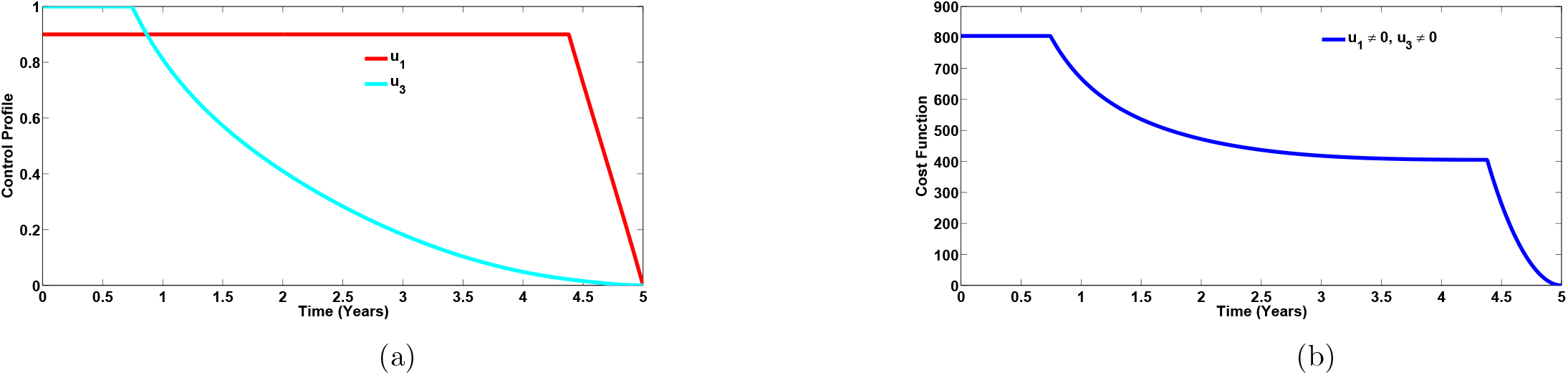
Control profile (Figure 6 (a)) and Cost function (Figure 6(b)) for the combined effects of the controls *u*_1_ and *u*_3_ on the dynamics of the HPV-Chlamydia trachomatis co-infection model (3). Here, *β*_HP_ = 1.35*, β*_CL_ = 1.0. All other parameters are as in Table 2

#### 5.2.3 Strategy C: Chlamydia trachomatis prevention (*u*_2_ ≠ 0) and treatment (*u*_4_ ≠ 0) controls

Implementing Chlamydia trachomatis-only intervention controls (Chlamydia trachomatis prevention (*u*_2_) and treatment (*u*_4_)), without applying any HPV intervention control (*u*_1_ = *u*_3_ = 0), it is observed in Figures 7(b) and 7(c), that the total number of individuals singly infected with Chlamydia trachomatis and the total number of individuals dually infected with HPV and Chlamydia trachomatis, respectively, are less than the populations when no control strategy is applied. In addition, this control strategy has an indirect benefit on the total number of individuals singly infected with HPV (see Figure 7(a)). Particularly, despite preventing 35,896 new Chlamydia trachomatis cases, this intervention strategy also prevents 6,970 new cases of HPV infections. Moreover, this intervention strategy averts 3,304 new co-infection cases. It is interesting to note that the Chlamydia trachomatis-only intervention strategy averts more co-infection cases compared to HPV-only intervention strategy (strategy B). The simulation results agree with the epidemioloical reports in [34, 35] that prior Chlamydia trachomatis infection increases susceptibility to multiple infections. The control profile for this strategy presented in Figure 8 (a) shows that control *u*_3_ is at its peak for the first 4.5 years and finally declines to zero. Similarly, the control *u*_4_ is at the peak level of 100% for the first 2.7 years before gradually declining to zero at final time (when *t* = 5 years). The cost function for the combined effects of these two controls is given by Figure 8 (b).

**Figure 7:**
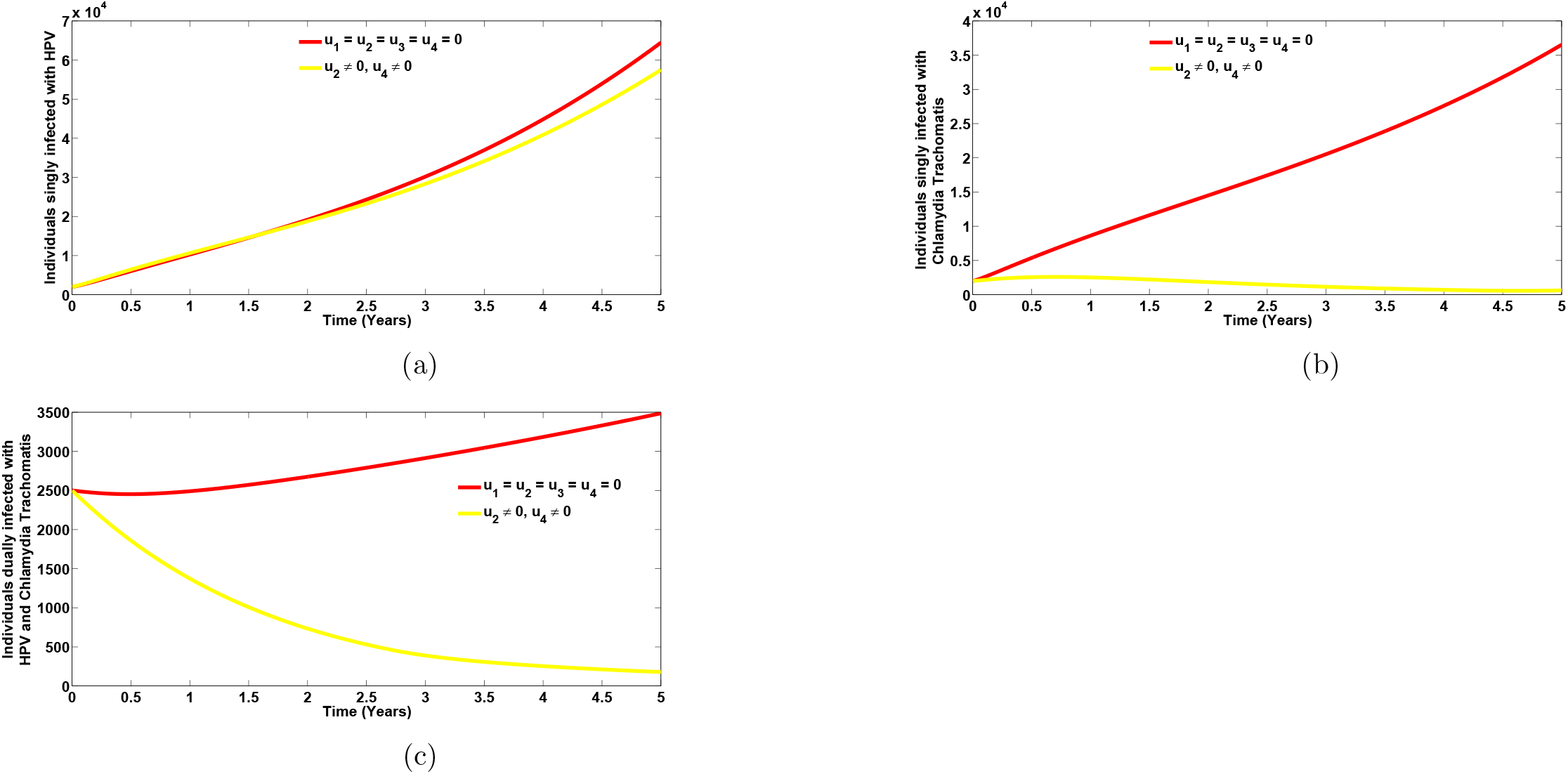
Plots of the total number of individuals singly infected with HPV (Figure 7 (a)), total number of individuals singly infected with Chlamydia trachomatis (Figure 7 (b)), as well as the total number of individuals dually infected with HPV and Chlamydia trachomatis (Figure 7 (c)), in the presence of Chlamydia tracahomatis prevention (*u*_2_ ≠ 0) and treatment (*u*_4_ ≠ 0) controls. Here, *β*_HP_ = 1.35*, β*_CL_ = 1.0. All other parameters are as in Table 2

**Figure 8:**
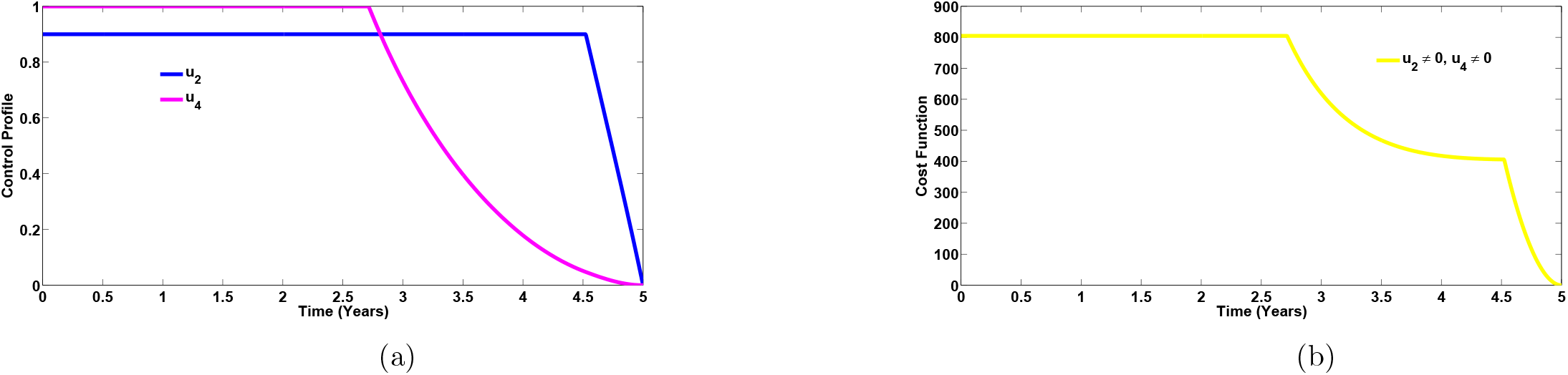
Control profile (Figure 8 (a)) and Cost function (Figure 8 (b)) for the combined effects of controls *u*_2_ and *u*_4_ on the dynamics of the HPV-Chlamydia trachomatis co-infection model (3). Here, *β*_HP_ = 1.35*, β*_CL_ = 1.0. All other parameters are as in Table 2

#### 5.2.4 Strategy D: HPV screening (*u*_1_ ≠ 0) and Chlamydia trachomatis treatment (*u*_2_ ≠ 0) controls

The plots of the total number of infected individuals when HPV screening (*u*_3_ ≠ 0) and Chlamydia trachomatis treatment (*u*_4_ ≠ 0) controls are applied, are given by Figures 9 (a) – 9 (c). It is observed that when this control strategy is administered, there is a significant reduction in the total number of individuals singly infected with HPV (Figure 9 (a)), total number of individuals singly infected with Chlamydia trachomatis (Figure 9b) and the total number of individuals dually infected with HPV and Chalmydia trachomatis (Figure 9 (c)). In particular, despite averting 29,150 new cases of HPV infections and preventing 13,600 new cases of Chlamydia trachomatis infection, strategy D also averts 2,606 new co-infection cases. The control profile for this strategy given in Figure 10 (a) shows that control *u*_3_ is at its peak for the first 2.7 years and ultimately declines to zero at final time (when *t* = 5 years). In a similar manner, the control *u*_4_ is at the maximum level of 100% for the first 4.0 years before finally declining to zero. The cost function for the combined effects of these two controls is given by Figure 10 (b).

**Figure 9:**
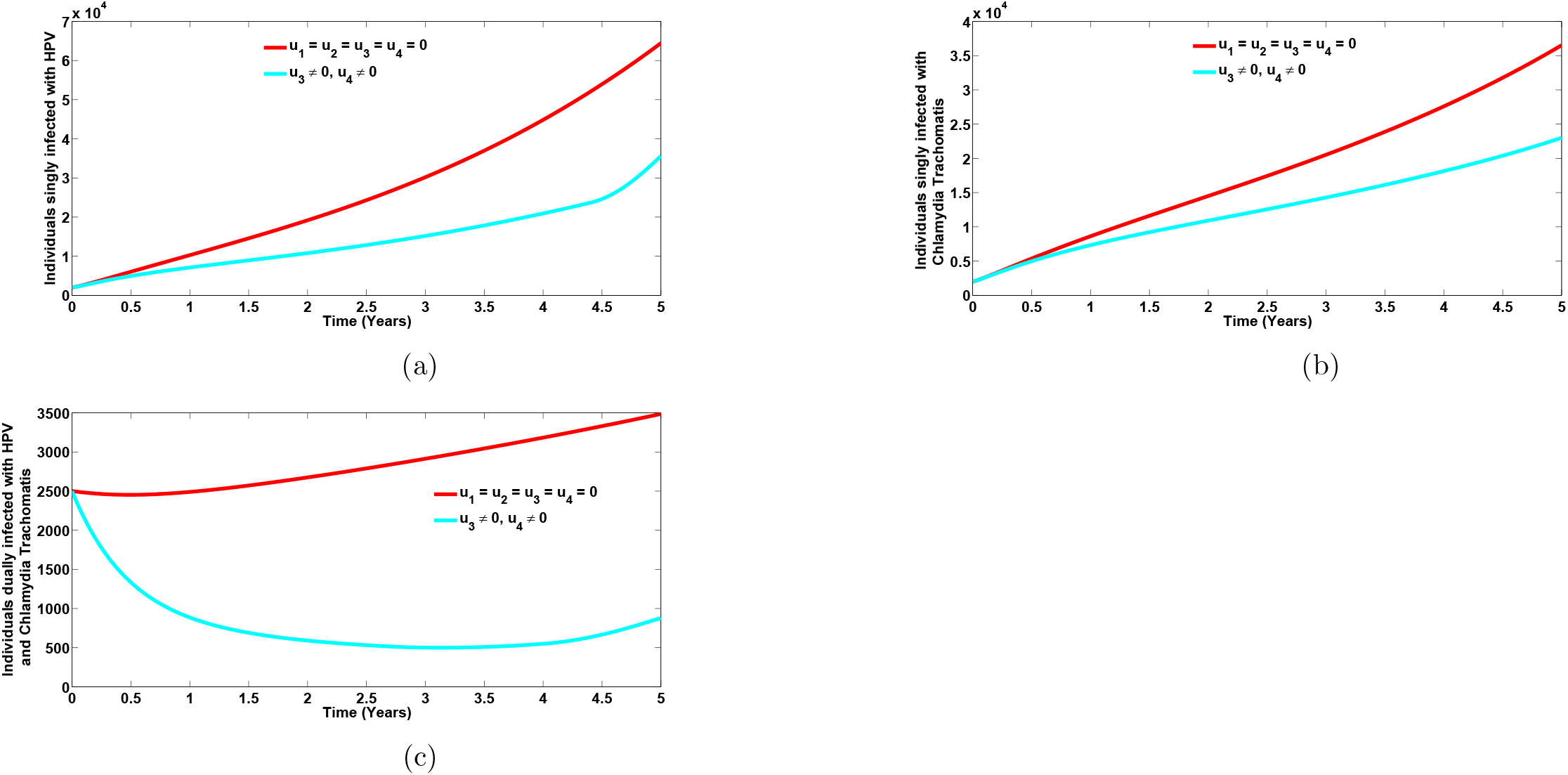
Plots of the total number of individuals singly infected with HPV (Figure 9 (a)), total number of individuals singly infected with Chlamydia trachomatis (Figure 9 (b)), as well as the total number of individuals dually infected with HPV and Chlamydia trachomatis (Figure 9 (c)), in the presence of HPV screening (*u*_3_ ≠ 0) and Chlamydia trachomatis treatment (*u*_4_ ≠ 0) controls. Here, *β*_HP_ = 1.35*, β*_CL_ = 1.0. All other parameters are as in Table 2

**Figure 10:**
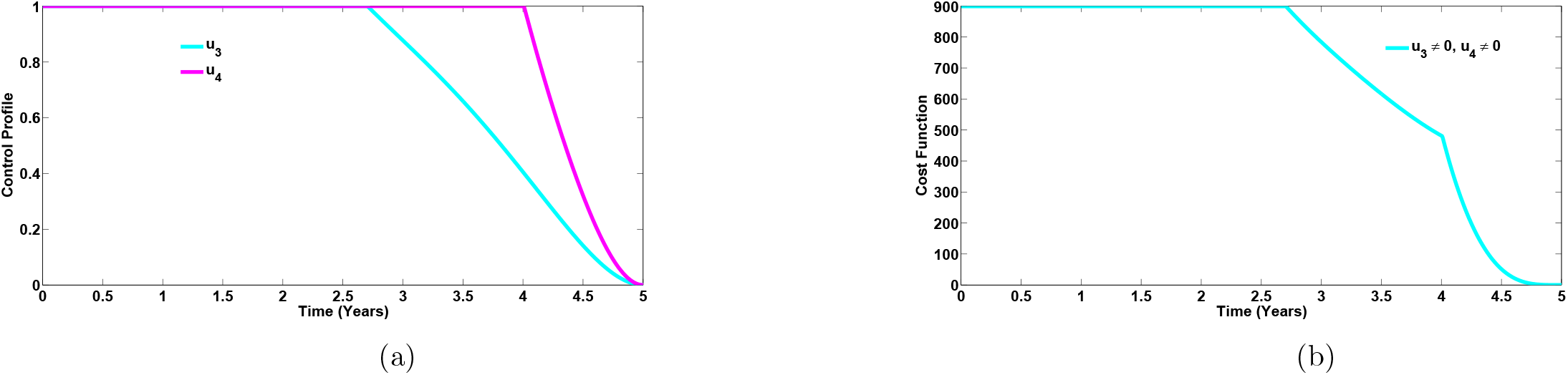
Control profile (Figure 10 (a)) and Cost function (Figure 10) for the combined effects of controls *u*_3_ and *u*_4_ on the dynamics of the HPV-Chlamydia trachomatis co-infection model (3). Here, *β*_HP_ = 1.35*, β*_CL_ = 1.0. All other parameters are as in Table 2

Simulations of the controls *u*_1_ and *u*_2_, where the weight constants *χ*_1_ and *χ*_2_ are varied, while fixing the values of the other weight constants *χ*_3_ = *χ*_4_ = 400, are depicted in Figures 11 (a) and 11 (b), respectively. It is observed from Figure 11 (a), that the control *u*_1_ was at its maximum value for 2 years, when *χ*_1_ = *χ*_2_ = 5000; was at its peak value for 3.7 years when *χ*_1_ = *χ*_2_ = 1000 and was at its highest value for 4.2 years when *χ*_1_ = *χ*_2_ = 500, respectively, before steadily declining to zero at final time (when *t* = 5 years). A similar trend is observed for the control profile of *u*_2_ when *χ*_1_ and *χ*_2_ are varied. It is seen from Figure 11 (b) that the peak of *u*_2_ lasted for 2.9 years, 4.4 years and 4.7 years, respectively, before declining steadily to its lower bound, when *χ*_1_ = *χ*_2_ = 5000, *χ*_1_ = *χ*_2_ = 1000 and *χ*_1_ = *χ*_2_ = 500, respectively. It is worthy of note that decreasing the weight constants from 5000 to 500 over time, increases the duration of the peak values of the controls *u*_1_ and *u*_2_ in minimizing the total number of infected individuals.

**Figure 11:**
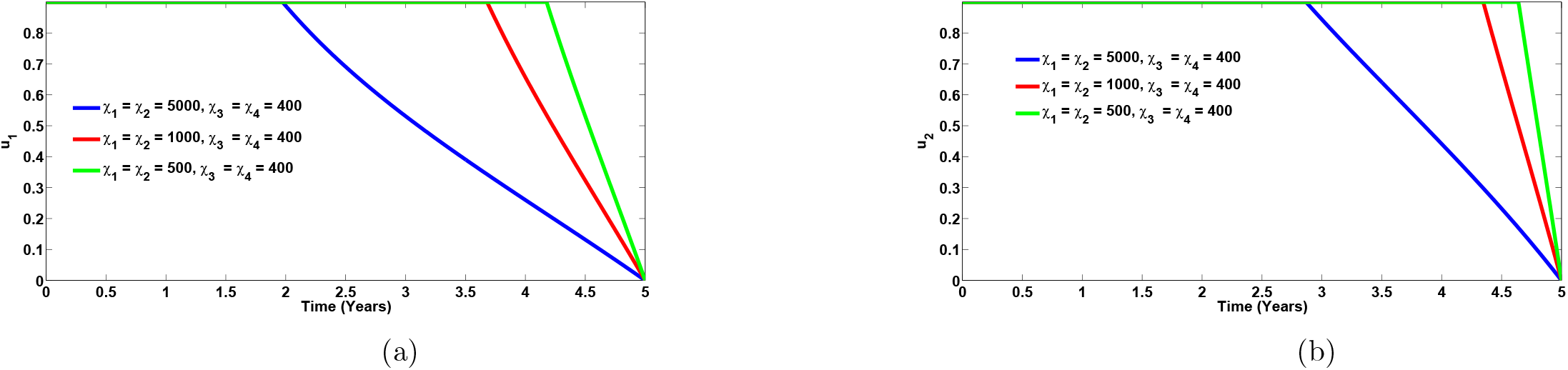
Plots of the controls *u*_1_ and *u*_2_ at different values of the weight constants χ_1_ and χ_2_. Here, *β*_HP_ = 1:35; *β*_CL_ = 1:0. All other parameters are as in Table 2

Also, the simulations of the controls *u*_3_ and *u*_4_, while keeping the weight constants *χ*_1_ and *χ*_2_ fixed and varying the values of *χ*_3_ and *χ*_4_, are depicted in Figures 12 (a) and 12 (b), respectively. It is observed from Figure 12 (a), that the control *u*_3_ was at its maximum value for 1.5 years, when *χ*_3_ = *χ*_4_ = 5000; was at its peak value for 3.4 years when *χ*_3_ = *χ*_4_ = 1000 and was at its highest value for 4.0 years when *χ*_3_ = *χ*_4_ = 400, respectively, before steadily declining to its lower bound when *t* = 5 years. A similar trend is observed for the control profile of *u*_4_ when *χ*_3_ and *χ*_4_ are varied. It is observed from Figure 12 (b) that the peak of *u*_4_ lasted for 1.5 years, 3.4 years and 4.0 years, respectively, before declining steadily to zero, when *χ*_3_ = *χ*_4_ = 5000, *χ*_3_ = *χ*_4_ = 1000 and *χ*_3_ = *χ*_4_ = 400, respectively. More so, It is equally observed that decreasing the weight constants from 5000 to 400 over time, increases the duration of the peak values of the controls *u*_3_ and *u*_4_ in minimizing the total number of infected individuals.

**Figure 12:**
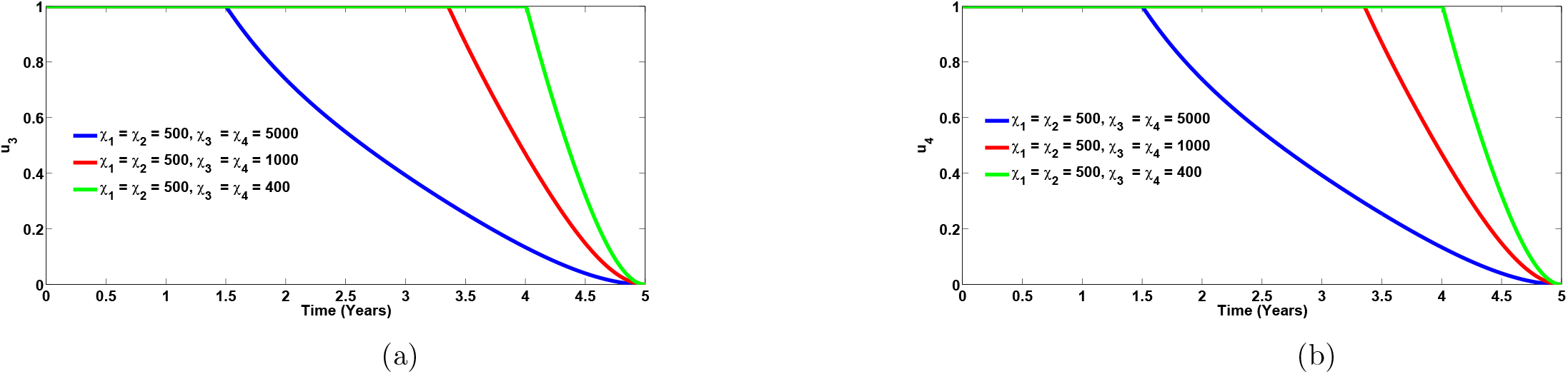
Plots of the controls *u*_3_ and *u*_4_ at different values of the weight constants *χ*_3_ and *χ*_4_. Here, *β*_HP_ = 1.35*, β*_CL_ = 1.0. All other parameters are as in Table 2

### 5.3 Cost-effectiveness analysis

The cost-effectiveness analysis is now applied to assess and evaluate the benefits associated with the health intervention strategies in order to justify the costs of the strategies [3]. This is obtained by relating the differences between the health outcomes and costs of those interventions, achieved by computing the incremental cost-effectiveness ratio (ICER), which is defined as the cost per health outcome. It is given by:

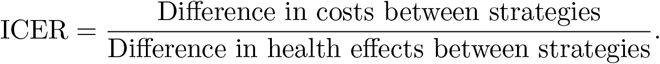

**Table 3:** Increasing order of the total infections averted due to the control various strategies.

| Strategy | Total infection averted | Total cost (\$) | ACER | ICER |
| --- | --- | --- | --- | --- |
| D: $u_3(t), u_4(t)$ | 45, 356 | 899.1 | 0.0198 | 0.0198 |
| C: $u_2(t), u_4(t)$ | 46, 170.4 | 809.8 | 0.00175 | -0.1097 |
| B: $u_1(t), u_3(t)$ | 73, 949 | 804.8 | 0.0109 | -0.000180 |
| A: $u_1(t), u_2(t)$ | 103, 157 | 804 | 0.00779 | -0.00002739 |

The total number of infections prevented and the total cost of the strategies applied are calculated in Table 3. The total number of infections averted is obtained by computing the difference between the total number of individuals when controls are administered and the total number when no control is applied. Likewise, the cost functions 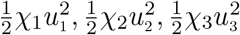 and 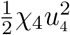, are applied over time, to compute the total cost for the various strategies implemented. The cost-effectiveness of strategy D (HPV screening and Chlamydia trachomatis treatment controls) and strategy C (Chlamydia trachomatis prevention and treatment controls) are now compared.

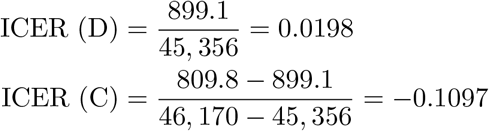

From ICER (D) and ICER (C), it is observed that the ICER for strategy D is greater than the ICER for strategy C. This implies that strategy D strongly dominates strategy C, indicating that strategy C is less costly and more effcetive in comparison with strategy D. As a result, strategy D is eliminated from subsequent ICER computations, as illustrated by Table 4. Strategies C and B are now compared.

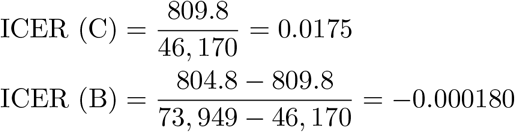

**Table 4:**
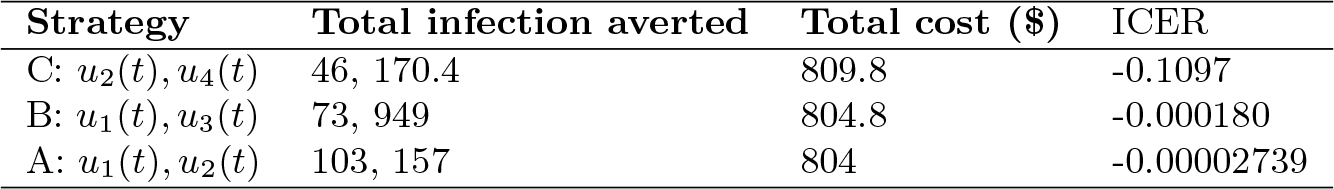
Increasing order of the total infections averted due to the control various strategies.

From ICER (C) and ICER (B), it is observed that a cost saving of 0.000180 is noticed for strategy B over strategy C. This implies that strategy C strongly dominates strategy B, indicating that strategy B is less costly and more effective in comparison with strategy C. Hence, strategy C is removed from subsequent ICER computations, as given by Table 5. Strategies B and A are now compared.

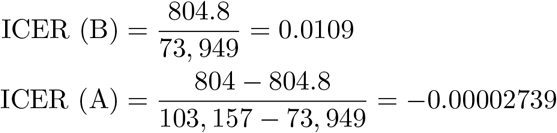

**Table 5:**
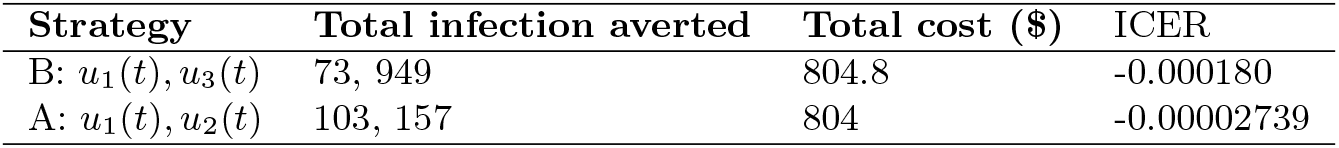
Increasing order of the total infections averted due to the control various strategies.

From ICER (B) and ICER (A), it is observed that strategy B strongly dominates strategy A, showing that strategy B is more costly and less effective in comparison with strategy A. In conclusion, strategy A (the strategy that implements HPV and Chlamydia trachomatis prevention controls) has the least ICER and is thus most cost-effective of all the control strategies in combating the co-infections of HPV and CHlamydia trachomatis. This result equally conforms with the results obtained before using the ACER method in Table 3, that strategy A is the most cost-effective strategy.

## 6 Conclusion

In this work, a co-infection model for human papillomavirus and Chlamydia trachomatis with cost-effectiveness optimal control analysis was developed and analyzed. Using the approach in Castillo-Chavez [4], the disease-free equilibrium of the model was proven not to be globally asymptotically stable. The model was shown to undergo the phenomenon of backward bifurcation when the associated reproduction number is less than unity. The HPV re-infection term (*ε*_P_ ≠ 0) induced the phenomenon of backward bifurcation in the HPV-Chlamydia trachomatis co-infection model. In addition, analysis of the HPV-only sub-model revealed the co-existence of two equilibria (a stable disease free equilibrium and a stable endemic equilibrium) when the reproduction number is less than unity. It was shown that HPV re-infection also induced the phenomenon of bakward bifurcation in the HPV-only sub-model. The necessary conditions for the existence of optimal control and the optimality system for the co-infection model was established using the Pontryagin’s Maximum Principle [30].

From the qualitative analysis of the model, it was observed that HPV re-infection *ε*_P_ ≠ 0, induced the phenomenon of backward bifurcation in the HPV-Chlamydia co-infection model. The epidemiological interpretation is that if recovery from HPV does not confer lifelong immunity, then the control of HPV-Chlamydia trachomatis becomes difficult in the population, even when the associated reproduction number 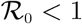. Hence, it is recomended that efforts should be made by government and health agencies to prevent re-infection with HPV so as to bring the burden of the co-infection of HPV and Chlamydia trachomatis very low at the community level. Moreover, sensitivity analysis of the model using the population of individuals co-infected with HPV and Chlamydia trachomatis revealed that the parameters that strongly influence the dynamics of the co-infection model are the effective contact rate for HPV transmissibility, *β*_HP_, the effective contact rate neccesary for Chlamydia trachomatis transmission, *β*_CL_, HPV screening rate for dually infected individuals, *η*_S1_, the modification parameters accounting for increased infectiousness of dually infected individuals, *φ*_P_(*φ*_L_), the modification parameter accounting for the infectiousness of individuals who have undergone HPV screening, *τ*_P_, and the modification parameters accounting for increased susceptibility to HPV (Chlamydia trachomatis) infection, 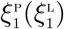. It is therefore, strongly recommended that efforts to bring down the burden of the co-infection of HPV and Chlamydia trachomatis should involve prevention strategies for both diseases, treatment and strict HPV screening policies.

Numerical simulations of the optimal control model showed that:

i. Focusing on HPV intervention strategies alone (HPV prevention and screening), in the absence of Chlamydia trachomatis control, a positive population level impact on the total number of individuals singly infected with Chlamydia trachomatis, is observed. This was shown in Figure 3 (b).
ii. Concentrating on Chlamydia trachomatis intervention controls alone (Chlamydia trachomatis prevention and treatment), in the absence of HPV intervention strategies, a positive population level impact on the total number of individuals singly infected with HPV, is observed. This was illustrated in Figure 7 (a).
iii. The strategy that combines and administers HPV and Chlamydia trachomatis prevention controls is the most cost-effective of all the control strategies in fighting the burden of the co-infection of HPV and Chlamydia trachomatis.

Furthermore, simulations of the various controls, where the weight constants are varied revealed that significant decrease in the weight constants from over time, increases the duration of the peak values of the associated controls in minimizing the total number of infected individuals (For instance, as observed in Figures 11 (a) and 11 (b), respectively).

## Data Availability

www.cdc.gov/std/chlamydia/stdfact-chlamydia-detailed.htm

